# Neuroimaging PheWAS and molecular phenotyping implicate *PSMC3* in Alzheimer’s Disease

**DOI:** 10.1101/2025.09.18.25336095

**Authors:** Xavier Bledsoe, Ting-Chen Wang, Yiyang Wu, Derek Archer, Hung Hsin Chen, Adam Naj, William S. Bush, Timothy J. Hohman, Logan Dumitrescu, Jennifer E. Below, Eric R. Gamazon

## Abstract

**INTRODUCTION:** Neuroimaging genetics have advanced Alzheimer’s disease (AD) research, yet frameworks mechanistically connecting genes to neurological outcomes via functional genomics are needed to elucidate genetic associations. To address this challenge, we assessed relationships between AD-associated variants and disease via their impact on gene expression and neuroimaging phenotypes.

**METHODS:** We mapped established AD genes to neuroimaging traits using NeuroimaGene atlas and predicted transcript-driven AD neurological features by comparing gene-derived neuroimaging features to clinical neuroimaging data. Genetic correlation and covariance analyses characterized shared genetic architecture between AD endophenotypes and neuroimaging features and identified neuroimaging features associated with dementia family history.

**RESULTS:** Our analyses implicate *PSMC3* expression as a strong contributor to AD pathophysiology and indicate AD endophenotypes, including dementia family history, linked to frontal cortex thickness, volume, and cerebrospinal fluid volume changes.

**DISCUSSION:** Our findings prioritize AD genes whose regulation is associated with vulnerable brain regions, offering a potential mechanistic framework for downstream functional validation.

## Background

Alzheimer’s disease (AD) is a complex polygenic neurodegenerative disorder with an estimated genetic heritability of 60-80%.^1,2^ The high heritability provides the opportunity to leverage genetic studies in characterizing the molecular basis of AD risk. Genome-wide association studies (GWAS) of AD-related phenotypes, including AD clinical diagnosis^3,4^, parental dementia status (AD by proxy)^5^, and rare variant analyses^6^ have all identified a host of genetic variants associated with AD risk. Investigating the functional implications of these variants is important in order to both advance our comprehension of the mechanisms underlying AD etiology and facilitate the development of innovative therapeutic interventions.

A range of approaches for integrating single-nucleotide polymorphism (SNP) and gene-level associations with neuroimaging data has emerged under the class of neuroimaging genomics. These approaches broadly aim to examine how genetic variation influences the structure and function of the brain in the context of disease. Prominent SNP-driven approaches include BrainXcan^7^, brain-wide GWAS^8^, and image-wide association studies (IWAS), both in the uni- and multivariate forms.^9,10^ However, the use of SNPs as predictor variables in these neuroimaging genetic studies presents limitations. One main challenge is that GWAS SNPs are often located within non-coding regions, limiting the ability to interpret their direct biological implications.^11^ This limitation underscores the utility of approaches like transcriptome-wide association study (TWAS). TWAS is a multivariate approach that utilizes expression quantitative trait loci (eQTL) data to identify associations between genetically regulated gene expression (GReX) and a trait of interest.^12^ Applied to AD, TWAS enables the mapping of noncoding risk SNPs to genes through GReX. These genes can then be aggregated as a transcriptomic signature and analyzed for mechanistic insight.^13,14^ While TWAS of AD and AD-associated phenotypes have been performed, the relationship of AD GReX to neuroimaging phenotypes remains an underexplored subject.

In this study, we investigated the relationship between molecular endophenotypes of AD and neuroimaging features **(Figure 1a, Supplementary Figure 1)**. The recently published NeuroimaGene resource used TWAS to identify associations between tissue-specific GReX and neuroimaging-derived phenotypes (NIDPs) generated from over 33,000 study participants in the UK Biobank (UKB).^15–18^ We thus leveraged NeuroimaGene to characterize the effect of AD TWAS genes on the structure and function of the brain **(Figure 1b)** using two analytical frameworks. First, we assessed for associations between AD TWAS genes and specific structural neuroimaging measures observed to differ between individuals with and without AD based on clinical studies.^19,20^ Secondly, we implemented an image-agnostic test to identify any brain regions whose structural morphology is associated with AD TWAS genes.^3^ To extend these analyses to SNP-level genetic endophenotypes, we also investigated local and genome-wide genetic correlations between brain morphology and established AD GWAS profiles for AD diagnosis and parental dementia **(Figure 1c)**. As a comparison, we also considered the phenotypic correlation investigating quantitative neuroimaging features associated with family history of dementia **(Figure 1d)** and clinical AD status **(Figure 1e)**. Details about each data source utilized in this study are provided in **Supplementary Table 1.** A strength of these analyses is that the neuroimaging measures implicated in our genetic analyses are derived from a set of largely healthy individuals who are younger than the typical AD patient. This mitigates imaging-based confounding introduced by aging and disease-mediated brain changes.^18^ Given the liability model of disease, we explored whether the mechanistic relationships between gene expression and brain structure apply to individuals irrespective of disease status.^21^ Our omics-informed neuroimaging analyses of genetic AD risk allow us to evaluate enrichment for clinically validated AD neuroimaging measures^20^ (**Figure 1f**). Through this multidisciplinary approach, we present a network of relationships linking diverse AD endophenotypes into an interconnected system of biological variables.

**Figure 1:**
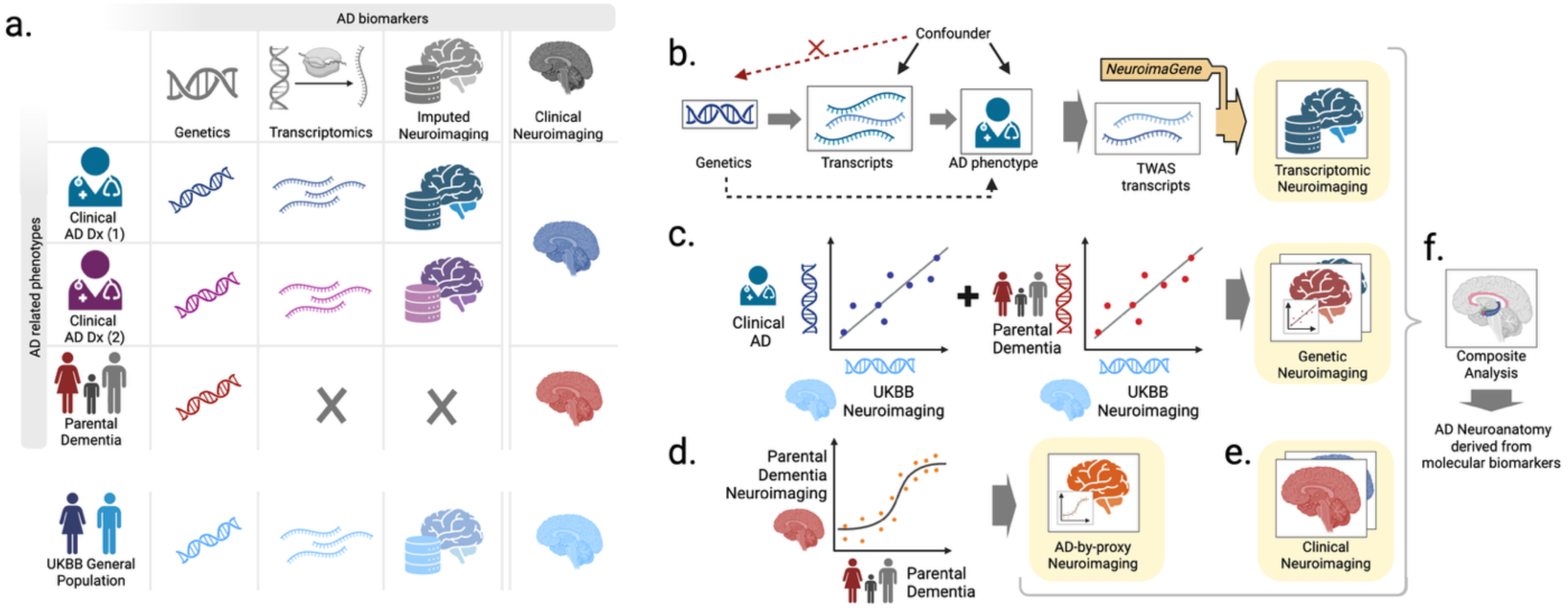
Schematic overview of analyses performed. **a.** Grid of main data resources used in the analysis. **b.** Directed acyclic graph of TWAS analyses with downstream imputation of neuroimaging features via NeuroimaGene. **c.** Visualization of genetic covariance analyses comparing the genetic architecture of clinical AD and parental AD with neuroimaging features. **d.** Logistic regression of neuroimaging feature measures and parental AD status. **e.** We leveraged clinical neuroimaging data associating brain features with AD status. **f.** We synthesized and compared the neuroimaging features obtained from transcriptomic, genetic, parental, and clinical approaches into a composite examination. UKBB – UK biobank; AD – Alzheimer’s Disease; Dx – Diagnosis; GWAS – Genome-wide association study; TWAS – Transcriptome-wide association study.

## Methods

### Genetic regulation of Alzheimer’s disease (AD) gene expression identification

From a recent study, we identified a set of genes with GReX associated with AD. Chen et al.^3^ reported 32 genes whose GReX is associated with AD disease status relative to healthy controls as determined in participants from the IGAP and ADGC consortium **(Supplementary Table 2).** The MetaXcan cross-tissue TWAS framework used gene expression models trained exclusively on brain tissues to determine these associations.^19^ Following causal inference via Mendelian randomization (MR) in their study, 23 of the gene-level associations retained statistical significance. We included GReX of these 23 genes as input variables for our image-agnostic and image-directed analyses documented below.

### AD-associated neuroimaging curation for image-directed analysis

Westman et al.^20^ used magnetic resonance imaging (MRI) scans of 699 subjects categorized as AD cases, controls, or those with mild cognitive impairment (MCI) from the Alzheimer’s Disease Neuroimaging Initiative to identify measures of the brain that differ between patients according to disease status **(Supplementary Table 3).** Unlike individuals in the UKB, these patients were recruited from the United States and Canada. They applied the FreeSurfer pipeline uniformly to scans from all patients to generate neuroimaging phenotypes corresponding to regional volume, cortical volume, surface area, and thickness. The atlases used for the parcellation of cortical and subcortical regions include the Destrieux atlas, the Desikan atlas, and the initial subcortical atlas reported in Fischl et al. 2002.^22–24^ Using these data, Westman et al.^20^ generated a prediction model trained on the neuroimaging measures for discriminating between patients with and without AD. The model includes the covariance of each neuroimaging measure with disease status as well as the confidence interval. We prioritized the 59 neuroimaging measures that demonstrate covariance with disease status and do not include zero within the confidence interval. The UKB includes neuroimaging measures derived from the same segmentation atlases as employed by Westman et al. Of note, the neuroimaging-derived phenotypes (NIDPs) in Westman et al.^20^ represent the average of the left and right hemispheres, while the UKB reports hemisphere-specific data. For each significant covarying NIDP in Westman et al., we selected the corresponding regions in both the left and right hemispheres aligned by the FreeSurfer segmentation atlas **(Supplementary Table 3)**. Control for multiple testing is described below. For this analysis, we utilized the NeuroimaGene resource described in Bledsoe et al.^16^ Of the 59 NIDPs reported by Westman et al., 57 are represented in the NeuroimaGene resource, with cortical measures duplicated in the left and right hemispheres.^16^ The absent phenotypes resulted from a lack of significant SNP-based heritability and failure of quality control in the neuroimaging phase of the UKB data generation. Including cortical regions duplicated in the right and left hemispheres, we selected a total of 110 of these NIDPs as AD-informative NIDPs for conducting image-directed analyses (**Figure 1e, Supplementary Table 4**).

### Multiple testing corrections for the image-directed analysis

Previous work demonstrates that gene expression is highly correlated and non-independent across tissue contexts.^25^ Therefore, we relied on the study-wide Benjamini-Hochberg False Discovery Rate (FDR) threshold of 0.05.^26^

### Gene expression in ROSMAP bulk RNAseq and AD endophenotypes

Processed bulk RNA sequencing (RNAseq) data from three brain tissues, dorsolateral prefrontal cortex (DLPFC, N=208), posterior cingulate cortex (PCC, N=490), and the head of the caudate nucleus (CN, N=673)^27^ was utilized to examine whether expression of genes from image-directed analyses with causal evidence is associated with AD endophenotypes, including AD pathology, amyloid beta (β) load (immunohistochemistry (IHC) staining), neurofibrillary tangles (IHC and silver staining), and cognitive function via performing multiple cross-sectional and longitudinal regression models (**Figure 2a-d, Supplementary Tables 5 & 6, Supplementary Figure 2)**. The cognitive function was represented as a global cognitive score derived from converting 19 cognitive tests raw scores to Z scores and then averaging them. The global cognitive score at the last visit pre-death was used for cross-sectional analyses. The longitudinal cognitive trajectories were derived from linear mixed-effects models estimating individual annual cognitive change. Covariates included age at death, sex, postmortem interval (PMI), and interval between last visit and death. For longitudinal models, time was modeled as the interval between a visit and the last visit, calculated in years. Statistical analyses were performed using R (version 4.2.1). Multiple testing correction was addressed by applying FDR per tissue and outcome across all genes tested.

**Figure 2:**
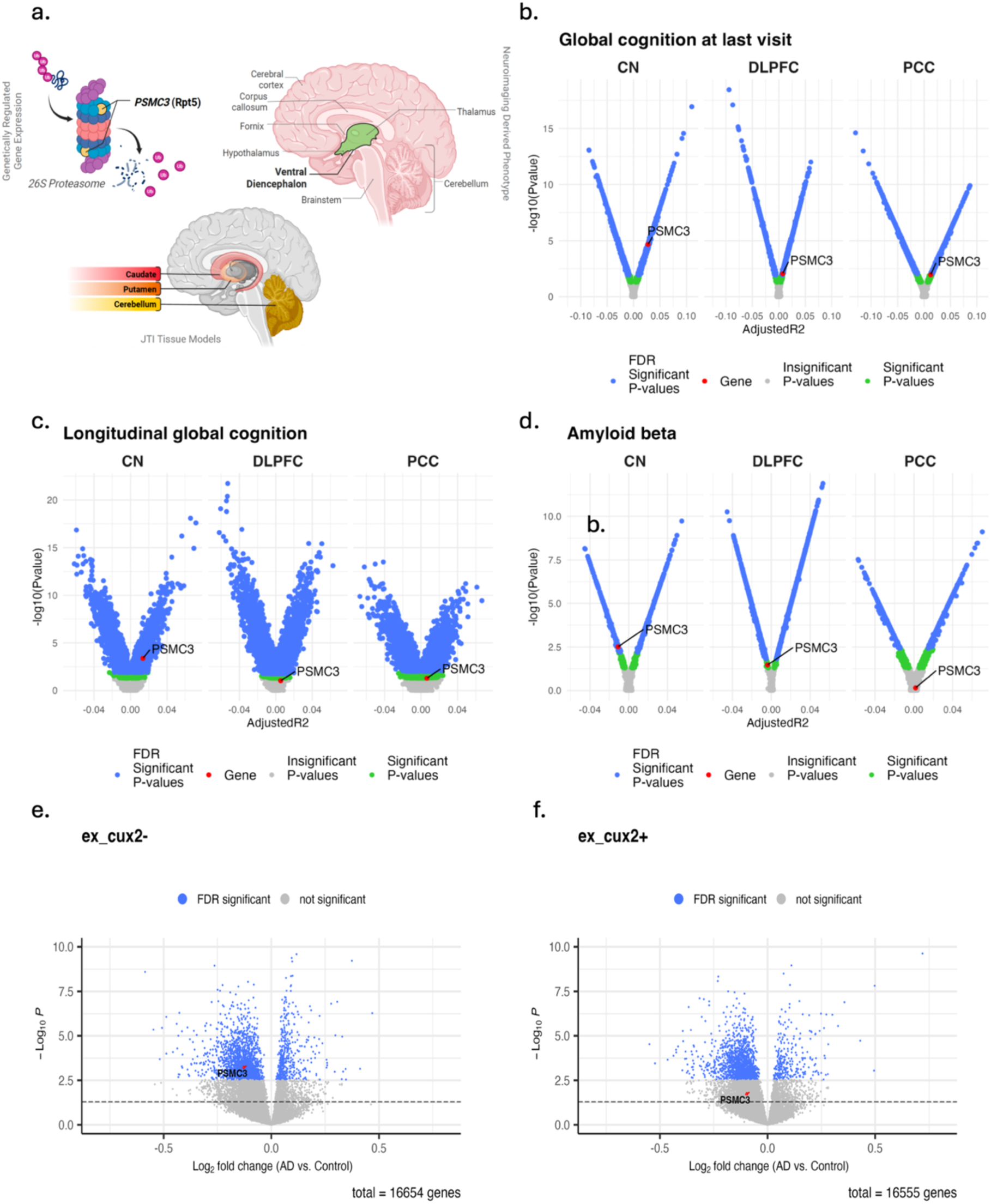
Visual summary of Mendelian randomization findings for *PSMC3* from image-directed analysis, corroborated by bulk and snRNA sequencing evidence from ROSMAP linking its expression to AD endophenotypes. **a.** GReX of *PSMC3* is associated with the volume of the ventral diencephalon according to 3 different gene expression models trained in the Caudate, Putamen, and Cerebellum. *PSMC3* encodes the protein Rpt5, which acts as an ATPase involved in substrate translocation in the 26S proteasome complex. *PSMC3* expression levels from bulk RNA-seq associated with **b.** global cognition score at last visit, **c.** longitudinal global cognition trajectory, and **d.** amyloid β deposit. The x-axis in the volcano plot represents the adjusted R^2^ value derived from the regression analyses. *PSMC3* expression levels from snRNA-seq associated with AD case/control status in **e.** cux2- and f. cux2+ excitatory neurons. The x-axis represents the log-fold change of *PSMC3* expression between AD cases and normal controls. The y-axis in **b-f.** represents the -log_10_ *P-*values for significance. The blue dots represent significant *P-*values (*P*_FDR_ < 0.05), the green dots represent nominally significant *P-*values, and the grey dots represent insignificant *P-*values. The red dots and arrows indicated *PSMC3.* CN - caudate nucleus; DLPFC - dorsolateral prefrontal cortex; PCC - posterior cingulate cortex.

### Gene expression in ROSMAP single-nucleus RNA-seq and AD endophenotypes

We further investigated cell-type-specific gene expression profiles linked to AD endophenotypes of genes with causal evidence from image-directed analyses by utilizing the single-nucleus RNA sequencing (snRNAseq) data derived from DLPFC brain specimens of 424 post-QC ROS/MAP participants (syn31512863).^28^ Eight major cell types, cux2+/cux2-excitatory neurons, inhibitory neurons, astrocytes, microglia, oligodendrocytes, oligodendrocyte precursor cells (OPCs), and endothelial cells were included in the analyses. Briefly, exclusion criteria applied to genes with expression in fewer than 10% of all cells and cells that had over 20,000 or fewer than 200 total RNA unique molecular identifiers (UMIs), or if more than 5% of their reads mapped to mitochondria. The gene count matrix was derived from UMI count data from the RNA assay, normalized and scaled by utilizing “sctransform” R package https://github.com/satijalab/sctransform). The NEBULA-HL method, implemented within the NEBULA R package (version 1.2.0)^29^, was carried out to perform negative binomial lognormal mixed models in a cell-type-specific manner on the snRNAseq data.^30^ Differential gene expression was assessed between participants with normal cognition (N= 142) and those with AD dementia (N=157) (**Figures 2e & f, Supplementary Table 7, Supplementary Figure 3**), with analyses covarying for age at death, sex, and PMI. These same covariates were included in modeling the associations between snRNAseq profile of genes and AD endophenotypes, including amyloid β load (IHC staining), neurofibrillary tangles density (IHC staining), and both cross-sectional and longitudinal cognitive function (**Supplementary Table 7, Supplementary Figures 4-7**). Statistical analyses were performed using R (version 4.2.1). Multiple testing correction was addressed by performing the FDR procedure per cell type within each tissue and outcome across genes tested.

### Image-agnostic analysis

For the image-agnostic analysis, we utilized the NeuroimaGene^16^ resource to examine associations between the previously described 23 AD-TWAS genes and cortical and subcortical NIDPs morphology as generated by the FIRST and FAST image segmentation protocols (**Figure 3, Supplementary Table 8, Supplementary Figure 8**).^31–33^ We opted for an atlas-wide FDR of 0.05 as our significance threshold. Next, we applied the Mendelian randomization joint tissue imputation pipeline (MR-JTI) to perform post hoc causal inference on these associations (**Supplementary Table 9**).^25^

**Figure 3:**
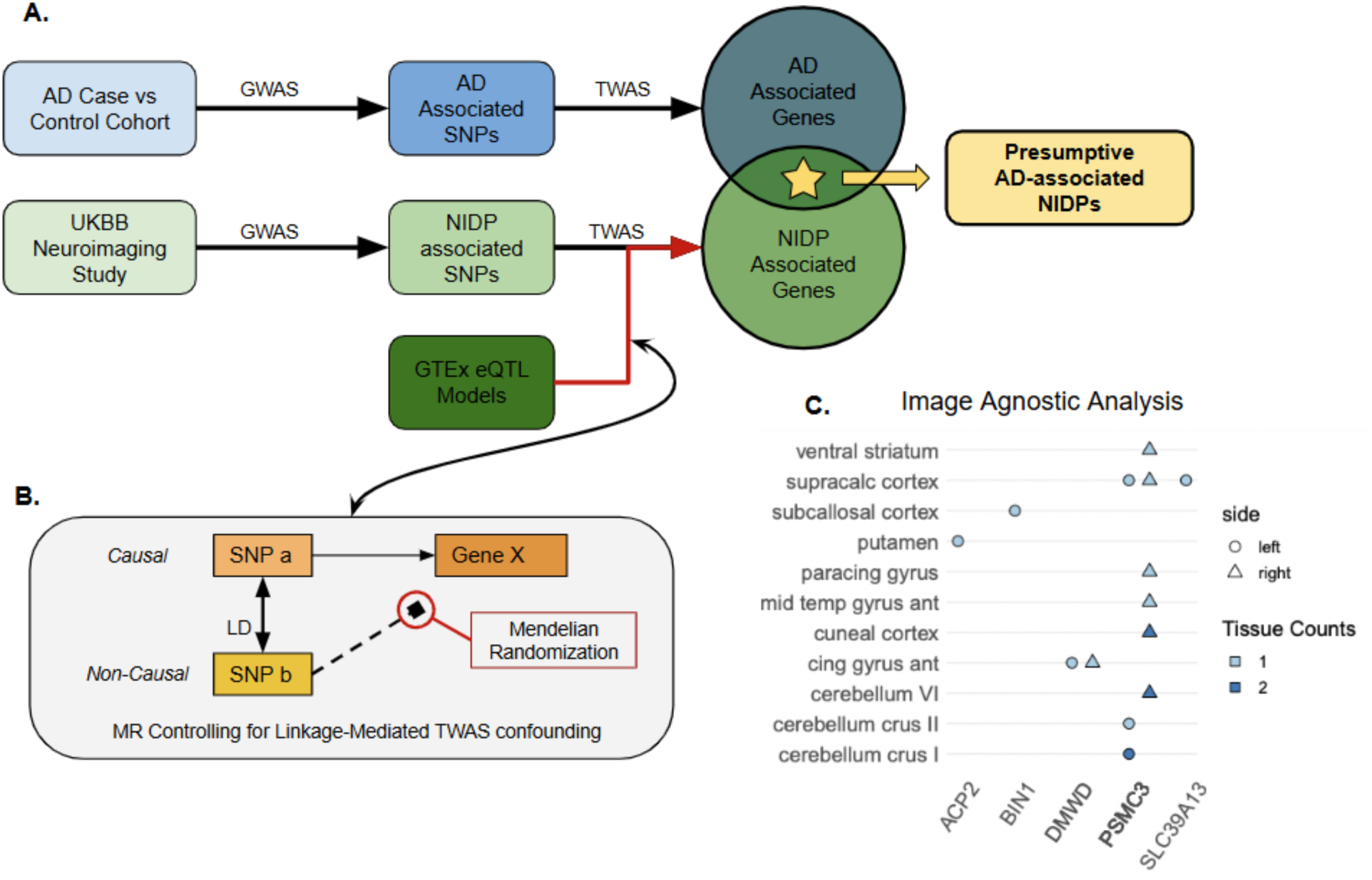
A. Schematic of TWAS intersectional methodology for the image agnostic approach. **B.** Schematic of horizontal pleiotropy mediated by linkage disequilibrium and the role of Mendelian Randomization in pleiotropy control. **C.** NeuroimaGene associations resulting from the image-agnostic analysis. Data points represent statistically significant associations between AD eGenes (x-axis) and NIDPs representing cortical and subcortical parcellations of the brain (y-axis). The color of each tile represents the number of tissue-specific gene expression models in which the eGene-neuroimaging association reached statistical significance in the MR-JTI causal inference analysis. The shape of the point specifies the lateral localization of the region to either the left or right hemisphere.

### Tissue context validation in the Human Protein Atlas

The Human Protein Atlas (HPA) provides bulk RNAseq data collected across 253 different tissues and spanning 19,023 genes.^34^ We accessed the data release that includes only information from HPA study participants instead of the consensus data to avoid sample overlap with GTEx. To validate the tissue contexts in which the AD-GReX associations were identified from the image-agnostic analysis, we first selected the genes from the association analysis on the full catalog of NIDPs that demonstrated significant causal effects on both AD and neuroimaging measures in MR. We then identified the tissue contexts in which these associations reached significance.^25^ We next mapped these tissues to those included in the HPA according to their neuroanatomic identifiers **(Supplementary Table 10)**. For each gene, we identified the measured expression across all 253 tissues from the HPA **(Supplementary Figures 9 & 10)**. We scored tissues on a sliding scale from 0 (lowest measured gene expression) to 1 (highest). We then ranked each tissue-gene pair according to the score of the tissue **(Supplementary Figure 11)**.

### Genetic correlation between family history of dementia and structural NIDPs

We identified loci associated with parental dementia from a GWAS by Marioni et al. from over three hundred thousand individuals from the UKB.^5^ We utilized LDSC to perform genetic correlation analysis between each NIDP in the UKB and the questionnaire-based parental dementia status using summary statistics (**Figure 4, Supplementary Table 11, Supplementary Figure 12**).^35^ LD scores were derived from the subset of European data from the 1000 Genomes Project.^36^ We retained NIDPs with nominally significant genetic correlations (p_uncorrected_ < 0.05).

**Figure 4:**
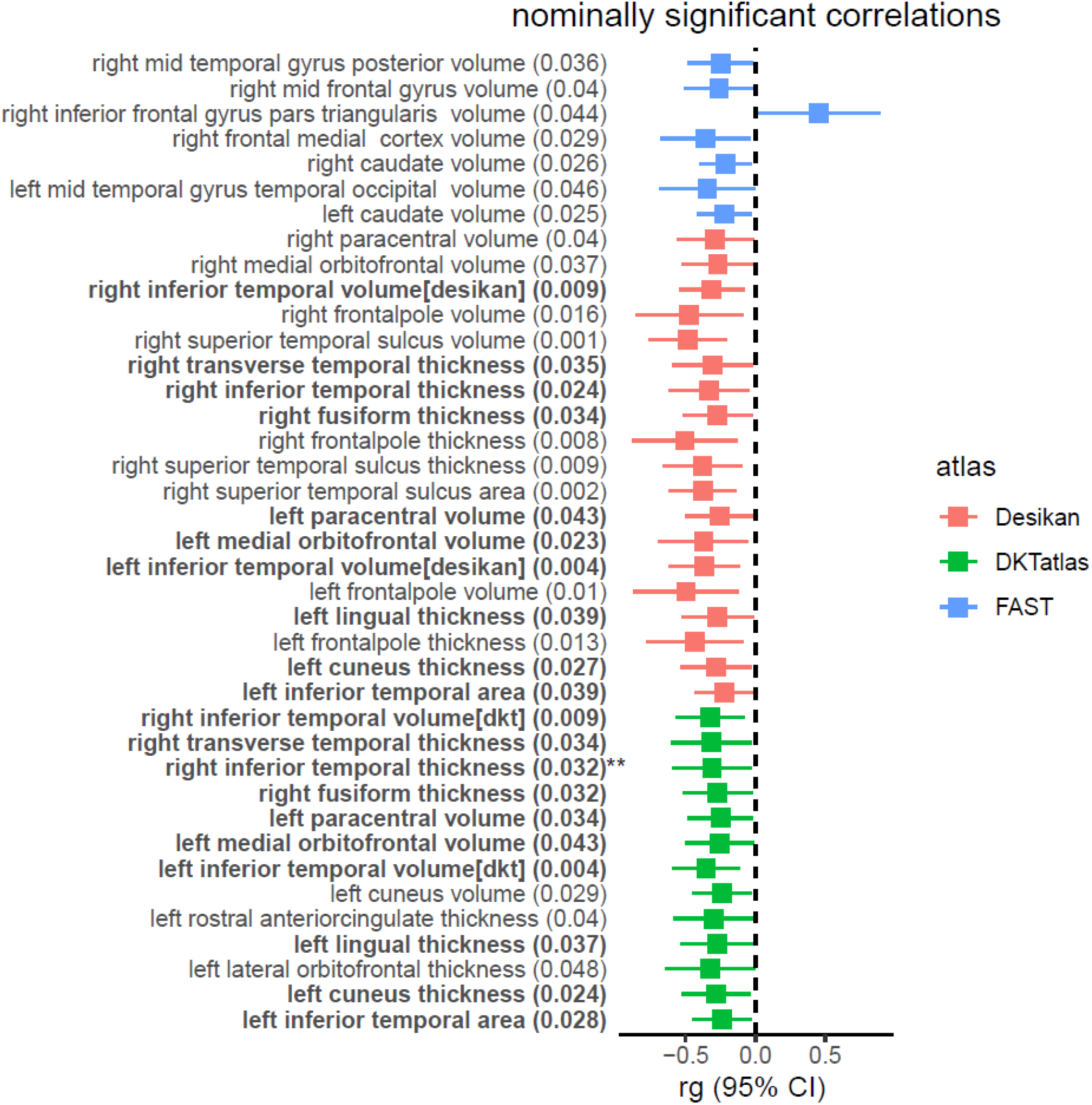
Nominally significant global genetic correlations between NIDPs and family history of dementia (p_uncorrected_ < 0.05). Genetic correlation was calculated using linkage disequilibrium score regression correlation analysis. Neuroimaging features that were significantly correlated with dementia family history in both the Desikan and DKT atlas are bolded. NIDPs that were significantly correlated with dementia family history and demonstrated significant covariance with AD status are marked with the double asterisk.

### Genetic covariance of AD status and structural NIDPs

We identified loci associated with clinical/pathology-confirmed AD from the GWAS by Kunkle et al. (N_cases_ = 35,274, N_controls_ = 59,163).^4^ We then performed genetic covariance analyses between 490 structural NIDPs, including those from the Desikan, DKT, and Fischl subcortical atlas, and AD status utilizing GeNetic cOVariance Analyzer (GNOVA).^37^ GNOVA is robust to sample overlap between GWAS datasets. Significance was defined as P_FDR_ < 0.05 **(Figure 5a, Supplementary Table 12, Supplementary Figure 13).** Additionally, we applied GNOVA to test for the genetic covariance between AD^4^ status and 46 NIDPs measured from the hippocampus, applying the same significance threshold as above **(Figure 5b, Supplementary Table 13).**

**Figure 5:**
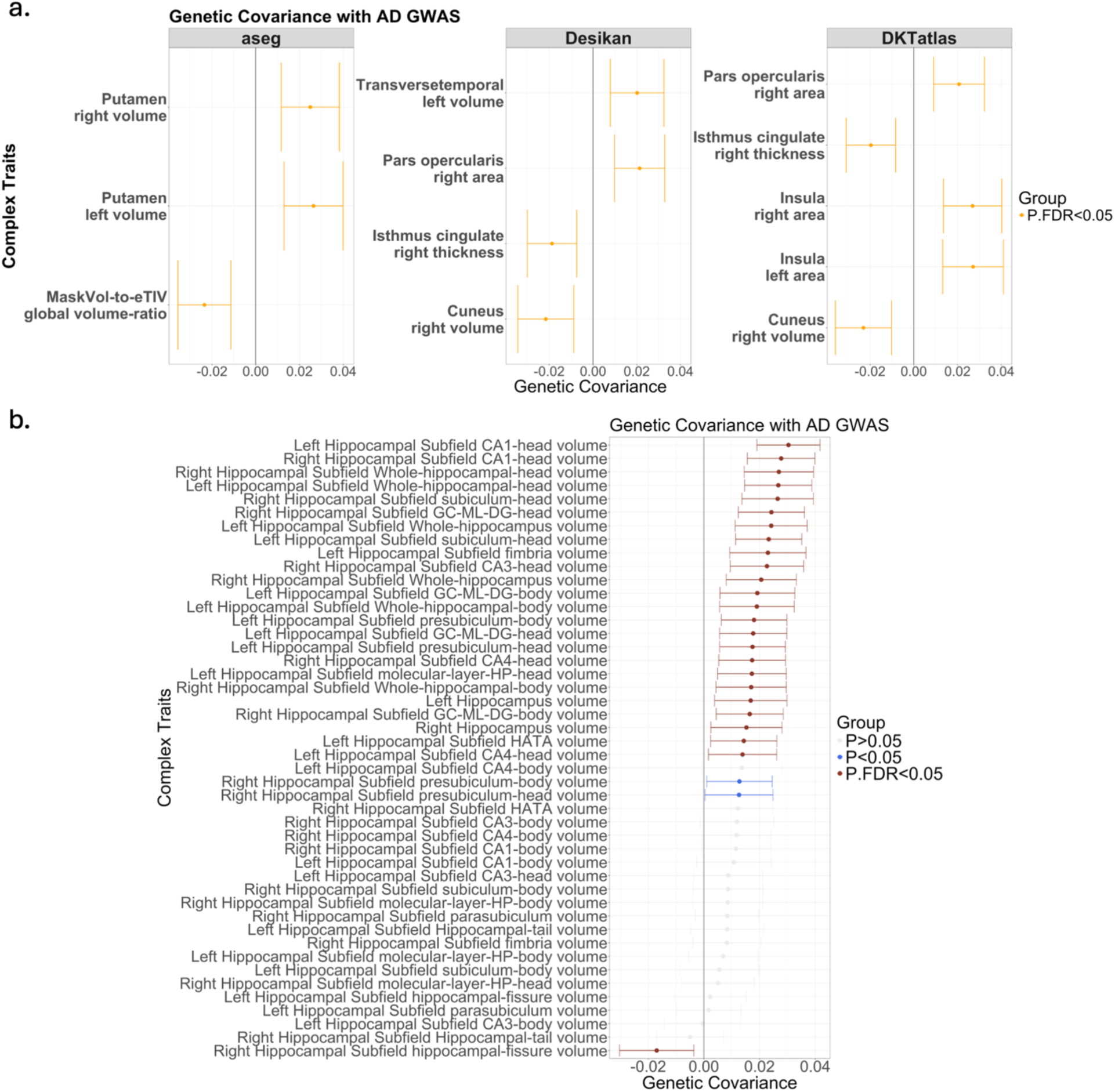
a. Genetic covariance calculated using GNOVA between NIDPs, processed using aseg, Desikan, and DKT atlases, and clinical AD status as indicated by the study of Kunkle et al. The x-axis depicts the estimates of genetic covariance. The y-axis represents the NIDPs. Each facet displays results from a different atlas. **b.** The genetic covariance results between hippocampus NIDPs and clinical AD status as indicated by Kunkle et al. The x-axis depicts the genetic covariance estimates, while the y-axis demonstrates the NIDPs. The significant results are shown in dark red, nominally significant results in light blue, and non-significant results in grey.

### Local genetic covariance of dementia family history and structural NIDPs

We investigated the local genetic covariance between parental AD status (using the Marioni et al.^5^ GWAS) and each previously implicated structural NIDP associated with GReX of previously implicated genes **(Supplementary Figures 14-22)**, using the SUPER GeNetic cOVariance Analyzer (SUPERGNOVA) tool.^38^ SUPERGNOVA leverages LDetect to partition the genome into local LD blocks for examining local genetic covariance among each GWAS pair.^39^ Significance was defined as the nominal SUPERGNOVA p < 0.05. In addition, we required the nominally significant genetic covariance signals to fall within 2 megabases upstream and downstream of the corresponding gene (based on genome build 37 coordinates).

### Phenotypic correlation between NIDPs and family history of dementia

We assessed the significance of empirical measurement differences in NIDPs across individuals with and without family histories of dementia in the UKB **(Supplementary Figure 23).** We first identified individuals with a family history of dementia according to the disease code 29626 (Alzheimer’s Disease) as occurring in either of the two data fields, “Illnesses of Father” and “Illnesses of Mother” **(Supplementary Table 14)**. Each individual was also tagged with data fields for sex, age at recruitment, and AD diagnosis to be used as covariates and filtering criteria. Among individuals self-reported either or both of their parents with AD dementia, we excluded those with neurologic diagnoses according to the presence of “G” category ICD codes as well as individuals with cerebrovascular accidents as noted by the “I6” prefix to ICD codes (**Supplementary Table 15**). Given the use of sex as a covariate, we excluded those individuals for whom the biological sex demonstrated a discrepancy with self-reported sex.

We first fit the NIDP data to a Weibull distribution using a Cullen and Frey chart. We then regressed the NIDP against family history of dementia with sex and age at recruitment as covariates. Using the Generalized Additive Models for Location Scale and Shape package in R, we performed this regression against a Weibull distribution: NIDP ∼ ADfamHx * Sex * ‘Age at recruitment’ **(Figure 6a, Supplementary Table 16).**^40^ NIDPs with distributions that did not match the Weibull distribution were evaluated according to a normal distribution **(Supplementary Table 16).** Nominally significant association of dementia family history with a NIDP was defined as p < 0.05 (**Figure 6b**). We repeated this analysis for the subset of NIDPs that demonstrated nominally significant correlations with dementia family history **(Figure 6c).**

**Figure 6:**
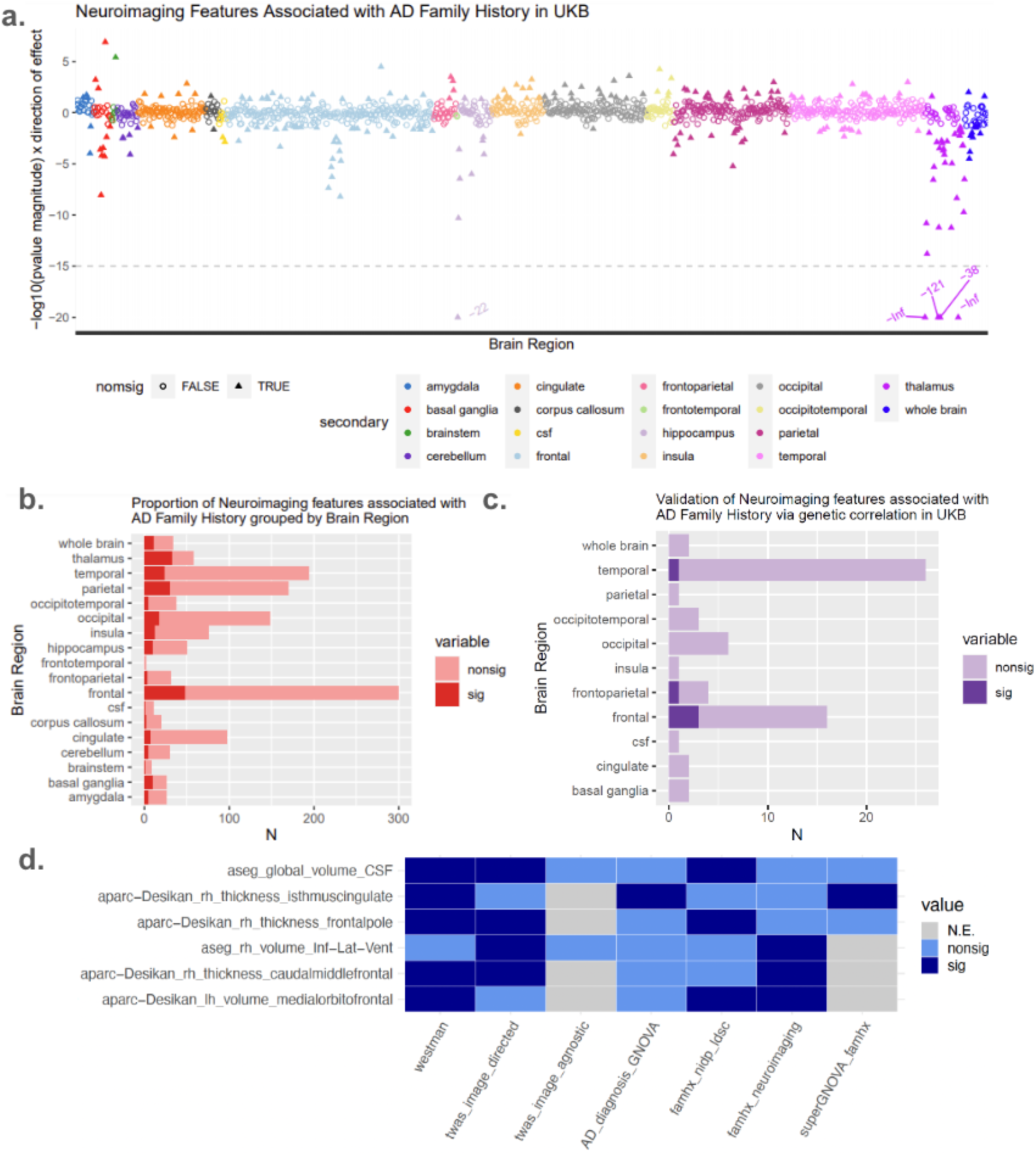
**a**. Neuroimaging features associated with dementia family history. The x-axis represents the surface area, volume, and thickness of cortical and subcortical regions as determined by the UKB. The y-axis represents the negative log of the effect size magnitude. This measure reflects the effect size between having a family history of dementia and the measurement of the region, adjusted for sex and age. Nominally significant findings (**p < 0.05**) are solid triangles while associations that did not pass significance are empty circles. Panels represent a division of findings according to effect size and direction of effect. Points are colored according to the neuroimaging atlas used to parcellate the brain regions in MRI protocols. **b.** We categorized each NIDP according to the named brain region it describes (y axis). The x-axis represents the number of imaging features. The solid red represents the nominally significant associations, while the pale red represents those associations that did not reach significance. **c.** Using the same named regions of the brain, we analyzed the subset of NIDPs that demonstrate nominally significant global genetic correlation with dementia family history. Of these phenotypes, those that demonstrated nominally significant size differences in people with dementia family history vs without are colored solid purple, while the lilac represents regions not observed to differ. **d.** Tile plot detailing neuroimaging features (x axis) that achieved nominally significant associations with at least two AD-related variables (y axis). Significant NIDP-phenotype associations are colored deep blue, while insignificant associations are light blue. Those associations that were not evaluated (N.E.) are shaded grey.

## Results

### *PSMC3* is an AD-associated gene that affects AD-associated NIDPs

We first sought to identify instances where transcriptomic profiles of AD-associated genes demonstrated statistically significant associations with AD-associated NIDPs in the UKB. We leveraged NeuroimaGene^16^, an atlas of associations of GReX with NIDPs, to further characterize genes previously identified in AD TWAS^3^ **(Supplementary Table 2)** through their associations with AD-associated NIDPs **(Supplementary Table 3).**^20^ Statistical significance was defined as a Benjamini-Hochberg FDR threshold of 0.05 **(Supplementary Table 4)**. We then assessed causality in observational data using Mendelian randomization joint tissue imputation (MR-JTI).^25^ Our assumption of causality depends on having genetic instruments (eQTLs) that are 1) strongly associated with GReX, 2) independent of confounders affecting both GReX and NIDPs, and 3) influence NIDPs only through GReX **(Supplementary Table 5)**.^25^ We identified a causal effect of *PSMC3* GReX on the volume of the ventral diencephalon in tissue models derived from the caudate, putamen, and cerebellum. The volume of the ventral diencephalon has been independently noted to differ in AD patients compared to healthy controls.^20^ The *PSMC3* gene encodes the ATPase 3 protein for the 26S proteasome involved in the digestion of proteins tagged for cellular clearance. *PSMC3* enables the full proteasome complex to cleave ubiquitinated peptides in an ATP-dependent manner **(Figure 2a)**. The ventral diencephalon, associated with *PSMC3* and implicated in both AD and AD neuroimaging, involves multiple subcortical structures, including the hypothalamus, mammillary body, subthalamic nuclei, substantia nigra, red nucleus, lateral geniculate nucleus (LGN), and medial geniculate nucleus (MGN).

### Transcriptome analysis implicates *PSMC3* in AD endophenotypes

To further investigate the relationship between *PSMC3* gene expression and AD pathophysiology, we performed several tests using ROSMAP bulk and single-nucleus RNA sequencing (snRNAseq) data (Methods). In bulk RNAseq analyses of the dorsolateral prefrontal cortex (DLPFC), posterior cingulate cortex (PCC), and caudate nucleus (CN), we identified three significant associations (*P*_FDR_ < 0.05) between *PSMC3* expression and AD endophenotypes. These include global cognition score at last visit (β=0.789, *P*_FDR_=4.91E-04, **Figure 2b**), longitudinal trajectory of global cognition (β=0.062, *P*_FDR_=3.90E-03, **Figure 2c**), and amyloid β depositions (β=-0.550, *P*_FDR_=3.24E-02, **Figure 2d**) – in the CN tissue. An additional significant association between *PSMC3* expression and global cognition at the last visit (β=0.513, *P*_FDR_=4.16E-02, **Figure 2b**) was observed in the DLPFC tissue. All bulk RNAseq results and additional visualizations are presented in **Supplementary Table 6 and Supplementary Figure 2**.

In the snRNAseq analyses of DLPFC, we observed a single significant association between *PSMC3* expression in cux2-excitatory neurons and AD diagnosis status (logFC=-0.114, *P*_FDR_=0.019, **Figure 2e**). All snRNAseq results and additional visualizations are available in **Supplementary Table 7, Supplementary Figures 3-7**.

### Atlas-wide association testing of AD-associated genes and cortical NIDPs

We next sought to identify NIDPs associated with AD-genes independently of clinical neuroimaging studies, referred to as image-agnostic analyses. We hypothesized that the genetic and transcriptomic drivers of AD may induce subtle changes in brain architecture that are overshadowed by the effects of accumulating amyloid β and tau pathologies later in the disease. As such, there may be important neuroimaging markers of AD risk that would be missed by restricting our analysis to clinically derived NIDPs taken from patients with developed AD. We conducted an unbiased examination of associations between GReX of AD-associated genes and T1-derived cortical and subcortical NIDPs from the FIRST and FAST^32^ cortical and subcortical segmentation protocols **(Supplementary Table 8)**, which have previously been used in neuroimaging studies of AD. For significant (P_FDR_ < 0.05) GReX-NIDP associations, we tested for causal effects using MR-JTI.^25^ This approach identified five AD-associated genes with putatively causal effects on 13 NIDPs **(Figure 3, Supplementary Table 9)**. Again, *PSMC3* shows the most GReX-NIDP associations, appearing significant in six tissues (P_FDR_ < 0.05). The GReX-NIDP associations for the remaining genes *ACP2*, *BIN1*, *DMWD*, and *SLC39A13* with causal evidence were significant in a single tissue **(Supplementary Figure 8).**

### Gene expression validation in the Human Protein Atlas

We next explored the tissue contexts of genes identified in our analyses. As such, we examined the tissue specificity of the significant genes identified in our NeuroimaGene image-agnostic analyses in a second RNAseq data source. The Human Protein Atlas (HPA) provides bulk RNAseq data collected across 253 different tissues and spanning 19,023 genes. Of the seven tissues represented in MR significant gene-tissue associations from our atlas-wide image-agnostic analysis, six tissues were available in the HPA (**Supplementary Table 10**). Four of the five genes from these MR significant associations showed evidence of causality in a single tissue. All four of these genes were expressed in the corresponding tissue in the HPA **(Supplementary Figures 9 & 10)**.

To evaluate the specificity of gene expression across the complete catalog of available tissues, all 253 tissues were systematically ranked based on their measured expression levels (Methods). Notably, *PSMC3* was observed to be expressed in the HPA in the same five discovery tissues **(Supplementary Figure 11)**. The *PSMC3* expression in the caudate nucleus ranks within the 95^th^ percentile among all 253 tissues in the HPA. This observation aligns with the *PSMC3* GReX associations identified through image-directed analyses in the caudate, putamen, and cerebellum, with the volume of the ventral diencephalon (**Supplementary Table 5**). Overall, the significance of the discovery tissues derived from the NeuroimaGene is substantially corroborated in a second, independent cohort.

### Genetic correlation between family history of dementia and structural NIDPs

Here we aimed to investigate the neurological implications of SNP-mediated AD risk among individuals with a familial history of dementia. To exclude the broad array of dementia risk factors inherited through non-genetic mechanisms, we calculated the genetic correlation between familial dementia history and various NIDPs. We hypothesized that the findings from this analysis would identify NIDPs associated with AD in clinical neuroimaging studies. Parental status was employed as a proxy for genetic AD risk. Genetic correlation analyses were performed utilizing GWAS summary statistics pertaining to parental history and cortical and subcortical NIDPs derived from the UKB. Nineteen neuroimaging measures from the Desikan Atlas showed nominal significance (p < 0.05) in correlation with parental dementia status, indicating a shared genetic architecture **(Supplementary Table 11).** These neuroimaging traits encompassed measures of surface area, volume, or cortical thickness across nine different regions within the left or right hemisphere **(Figure 4, Supplementary Figure 12)**. Several of these cortical regions have been associated with prodromal AD according to Braak staging of both amyloid β and tau progression.^43^

### Genetic covariance of AD status and structural NIDPs

We next assessed the genetic covariance between AD diagnosis status and structural NIDPs processed using Fischl subcortical, Desikan, and DKT atlases, as well as the NIDPs measured from the hippocampus. We leveraged summary statistics from the GWAS by Kunkle et al. to characterize the genetic covariance of AD and structural NIDPs.^4^ We identified significant (*P*_FDR_ < 0.05) genetic covariance between AD diagnosis status and the volume of the cuneus, the thickness of the cingulate gyrus (isthmus), and the surface area of the right pars opercularis across the Desikan and DKT atlas. Additionally, we identified significant genetic covariance (*P*_FDR_ < 0.05) in the right and left surface area of the insula from the DKTatlas and the volume of the left transverse temporal gyrus from the Desikan atlas. The volume of the putamen on both hemispheres reached statistical significance **(Figure 5a, Supplementary Table 12, Supplementary Figure 13)**.

Regarding the hippocampal findings, 25 of the 46 subfields (54%) demonstrated significant (*P*_FDR_ < 0.05) genetic covariance with AD status (**Figure 5b, Supplementary Table 13**). The subcortical features with the greatest genetic covariance measures include the bilateral heads of cornu ammonis (CA) region 1, bilateral measures of the whole hippocampal head, and lastly, the volume of the right head of the subiculum.

### Local genetic covariance of parental dementia and structural NIDPs

The TWAS of parental dementia and genetic covariance analysis converged on two nominally significant overlapping associations **(Supplementary Table 17, Supplementary Figures 14-22)**. These two results were the associations between the *MS4A4E* expression level and the cortical thickness of the pars orbitalis and isthmus cingulate regions in the right hemisphere from the Desikan atlas **(Supplementary Table 17)**. *MS4A4E* is a membrane spanning protein that has been previously associated with AD and cerebral amyloid angiopathy.^44,45^

### Phenotypic correlation between NIDPs and family history of dementia

We used multivariate linear regression to examine the relationship between family history of dementia and T1 structural NIDPs **(Figure 6a, Supplementary Table 16)**. We identified 229 NIDPs with a nominally significant association (p_uncorrected_ < 0.05) with dementia family history, and 31 of these associations surpassed the Bonferroni significance threshold. Of the 31 Bonferroni significant associations, the majority described the thalamus, followed by the frontal cortex, the hippocampus, and the basal ganglia. Overall, 94% of these structural measures demonstrated changes consistent with atrophy (reductions in volume or increases in surface area) (**Supplementary Table 16)**.

## Discussion

Several studies have sought to identify neuroimaging and gene expression correlates of AD.^3,10,46–48^ Here, we performed analyses leveraging large extant resources to integrate genetic risk factors of AD, genetically determined gene expression, and genetic predictors of NIDPs to extend mechanistic and functional understanding of AD risk. We presented substantial evidence linking the expression of known AD risk genes with the morphology of specific brain regions known to be altered in patients with the disease. We used genetic covariance to show that either a parental history of dementia or a diagnosis of the disease is associated with changes in neuromorphology that can be detected via MRI.

Among the AD-related genes, *PSMC3* showed notable links with AD endophenotypes. The crucial role of *PSMC3* in effective intracellular protein degradation is of particular interest given the role of atypical protein aggregates in AD pathophysiology. Our Mendelian randomization (MR) analyses support a potential causal connection between *PSMC3* expression and multiple quantitative measures of brain morphology in healthy patients identified through both image-directed and image-agnostic methods (**Supplementary Tables 5 and 9**). Results from the ROSMAP bulk RNA sequencing identified significant positive associations between *PSMC3* expression and overall cognition at both cross-sectional and longitudinal timepoints, implying that reduced *PSMC3* levels in the CN and DLPFC tissues correlate with worse cognitive performance (**Supplementary Table 6**). Additionally, a negative association was found between *PSMC3* expression and amyloid β accumulation in CN, implying that increased *PSMC3* expression may confer protection against amyloid β buildup (**Supplementary Table 6**). Single-nucleus RNAseq in ROSMAP identified a single significant inverse relationship between *PSMC3* expression and AD diagnosis, specifically in cux2-excitatory neurons of the DLPFC, indicating observed dysregulation of *PSMC3* in AD patients compared to controls (**Supplementary Table 7**).

An additional notable finding is the significant causal association (from the MR analysis) between *PSMC3* in the caudate tissue and the volume of ventral diencephalon (DC), which is noted to be different in AD patients versus normal controls (**Supplementary Table 5**).^20,49,50^ Coupled with evidence of protective effects against amyloid β deposition and positive cognitive outcomes observed via its gene expression in the caudate nucleus of ROSMAP participants, this provides additional supporting evidence for *PSMC3’s* role in AD risk. Overall, our approach highlights how linking *PSMC3* GReX with NIDPs may inform hypotheses regarding brain regions influenced by *PSMC3* expression in relation to AD risk.

Synthesizing the neuroimaging findings from the genetic, transcriptomic, and clinical analyses, the top NIDPs implicated several regions with previous evidence for roles in AD **(Figure 6d, Supplementary Table 18)**. The increase of cerebrospinal fluid (CSF) volume in response to global cortical atrophy is our top finding and a well-documented association with AD.^51^ Additional outside work highlights the atrophy of the right inferior temporal cortex as causal in AD, a finding which we replicated in genetic covariance analyses of both AD status and parental dementia.^10^ Our third finding highlights the thickness of the isthmus in the right cingulate gyrus. Two recent publications highlight associations between the white matter architecture of the cingulate bundle with memory and cognitive performance as well as AD risk genes and AD polygenic risk scores.^52,53^

Of note, we implicated several previous findings of atrophy in the hippocampi. ^54,55^ Zammit et al.^54^ examined the relationship between hippocampal subfield morphology and cognitive performance as measured through figure and verbal memory testing in healthy adults. They identified statistically significant associations between volume of CA1, the subiculum, and the whole hippocampus with their cognitive measures. Multiple additional studies have reported associations between the subiculum and CA1 with AD. ^56–58^ The subiculum, CA1, and the whole hippocampus subregions in both hemispheres constitute six target subregions for our analysis. All six occur within the top seven results of our genetic covariance analysis, ranked by either p-value or covariance **(Figure 5b, Supplementary Table 13)**. Hippocampal atrophy is an early finding in AD and it has been hypothesized that these changes may reflect the region’s increased sensitivity to toxins such as amyloid β.^59^ Together, these data suggest that the genetic architecture of these hippocampal subregions itself is associated with AD and, relying on Zammit et al., that the specific subregions are involved in verbal and visual recall.

In evaluating the drivers of neurologic morphology, we considered the effects of gene expression rather than individual SNPs. The interpretation of gene expression is generalizable across populations in a way that SNP variation is not. Because we performed this work in a large population that is largely free of overt AD, our results reveal associations between gene expression and neuromorphology, i.e., inherited molecular changes that precede AD. The liability model of disease holds that risk is manifested through an accumulation of factors that drive a patient toward the upper end of a risk distribution. These same risk factors are distributed broadly across the population, usually at insufficient doses to mediate observable symptoms. The data presented here suggest that genetic and transcriptomic risk factors for AD result in subclinical biochemical and biological consequences that can be observed outside of the context of overt disease. Nevertheless, despite the convergence of neuroimaging findings in genetic risk, transcriptomic risk, and epidemiologic risk with the actual diagnosis, it is possible that over- or underexpression of gene products may have different consequences in the context of disease relative to states of health.

Several additional limitations merit discussion. Our analyses assessed only the effects mediated through gene expression. Future studies on downstream molecular traits (e.g., protein or metabolite) are warranted. Regarding data quality, the dementia family history is assessed via a self-report of familial history of dementia. While current evidence supports using this measure as a rough proxy for AD risk^60,61^, the diagnostic accuracy is likely to be reduced relative to a clinically phenotyped cohort.

We encountered several unexpected findings. Multiple NIDPs demonstrated genetic covariance with family history of dementia, but these were nearly all different regions from those linked with AD through the mechanism of GReX. Genetic covariance, in contrast to eQTL-based analysis, does not evaluate gene regulatory effects. We suspect that this discrepancy is primarily the result of parental dementia and positive AD status being different phenotypes. While the family history of dementia may predispose individuals to neuropathophysiological development, the regions impacted appear to differ from those influenced by the expression of AD-associated genes.

In summary, analysis of genetically determined expression of AD-associated risk genes provides a window into potential neurological mechanisms by which AD-associated variation confers disease risk. We showed that expression of *PSMC3* is associated with AD endophenotypes, including NIDPs, cross-sectional and longitudinal cognitive function, and amyloid β burden. Notably, in non-AD patients, GReX of *PSMC3* leads to similar neuroimaging changes, including the volume of ventral DC, as those observed in individuals with AD. Additionally, we demonstrated that family history of dementia and AD status are both associated with neuromorphology via genetic covariance, albeit different regions. Lastly, we presented the strongest composite support for endogenous influences on the frontal cortex, the global CSF volume, and the isthmus of the cingulate gyrus by AD-predisposing genetic variation. By integrating multiple layers of functional evidence, we provide a prioritization schema for identifying and contextualizing biochemical entities as areas of focus for future studies.

## Data Availability

All data produced in the present study are available upon reasonable request to the authors.

## Data availability

The bulk RNA sequencing data from 3 brain regions: the dorsolateral prefrontal cortex (DLPFC), posterior cingulate cortex (PCC), and the head of the caudate nucleus (CN) are available on the AMP-AD Knowledge Portal (syn23650893). The single-nucleus RNA sequencing data derived from 424 post-QC ROSMAP participants are available at Synapse (https://www.synapse.org/Synapse:syn31512863). The DOI for this dataset is https://doi.org/10.1038/s41588-024-01685-y.

## FUNDING and Acknowledgments

We acknowledge support from the following National Institutes of Health (NIH) grants: NIA R01AG073439, NIA R01AG061351, NHGRI R35HG010718, NHGRI R01HG011138, NIA AG068026, and NIGMS R01GM140287. We are grateful for funding from the Scott Hamilton CARES Foundation. ROSMAP data was obtained from the AD Knowledge Portal (https://adknowledgeportal.synapse.org), and the data collection was supported through NIA grants: P30AG010161 (ROS), P30AG072975, R01AG015819 (ROSMAP; genomics and RNAseq), R01AG017917 (MAP), R01AG036836 (RNAseq), and RF1AG057473 (single nucleus RNAseq).

## Consent Statement

All participants provided informed consent in their respective cohort studies.

