## Supplementary Figures for "Neuroimaging PheWAS and molecular phenotyping implicate *PSMC3* in Alzheimer’s Disease"

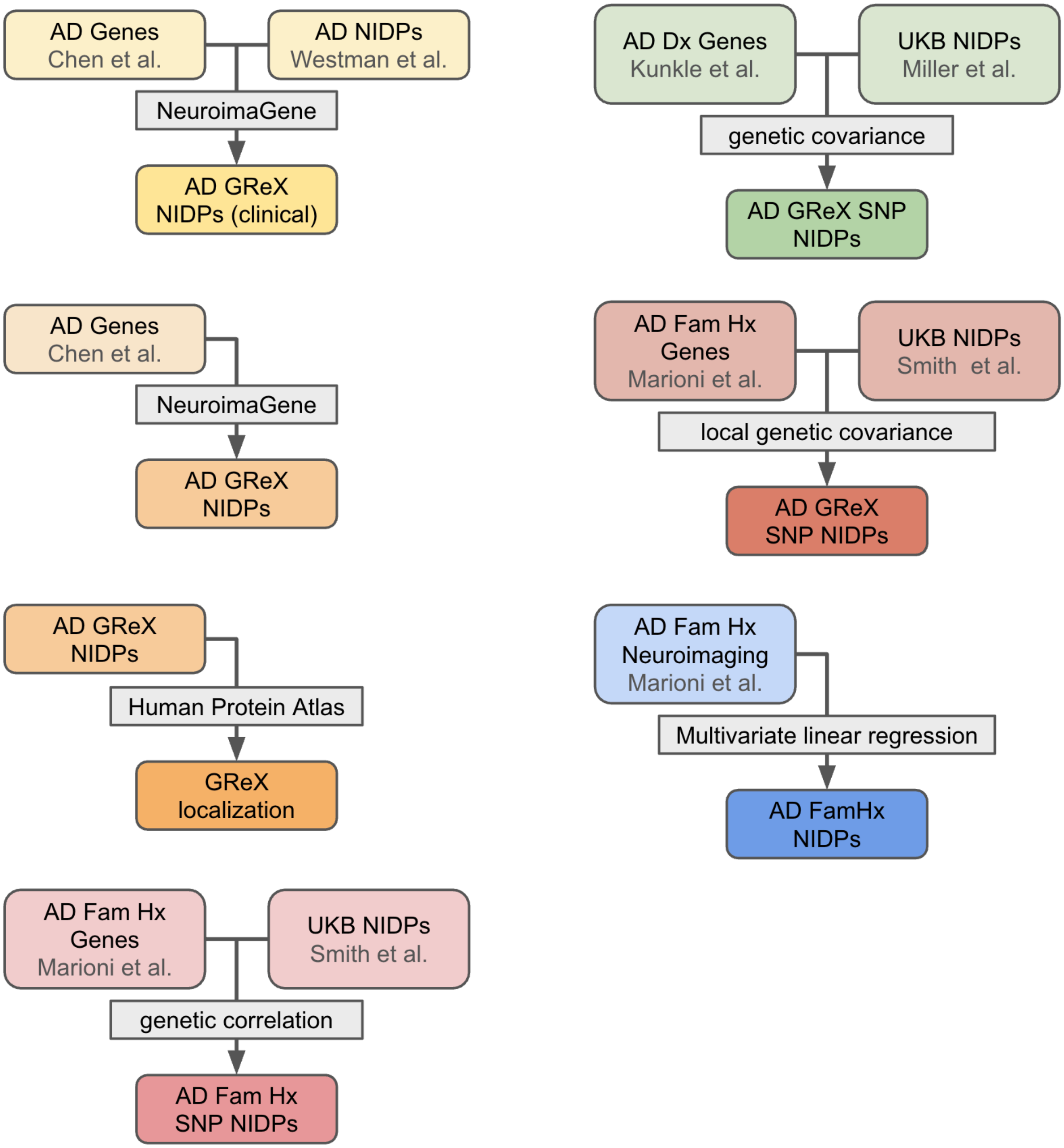


Figure S1: Overview of all analyses performed in this paper with resources and results in color and primary analytical method indicated in greyscale. AD – Alzheimer's Disease; NIDP – Neuroimaging derived Phenotype; GReX – genetically regulated gene expression; Fam Hx – family history; SNP – single nucleotide polymorphism; Dx – diagnosis

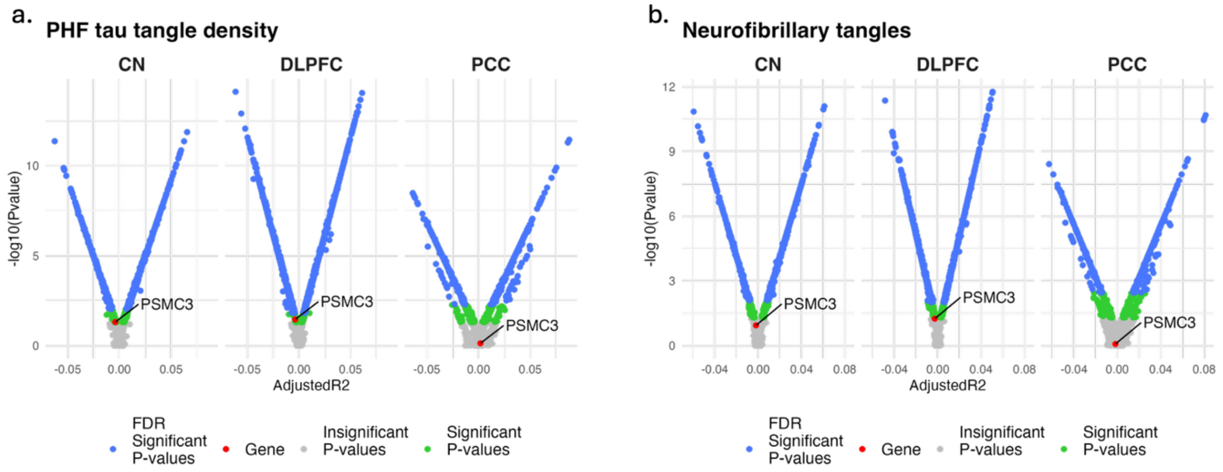


Figure S2: *PSMC3* expression levels from bulk RNA sequencing in ROSMAP associated with AD tau pathology assessed by (a) immunohistochemistry staining and (b) silver staining. The x-axis in the volcano plot represents the adjusted $R^2$ value derived from the regression analyses. The y-axis represents the $-\log_{10}$ *P*-values for significance. The blue dots represent FDR significant *P*-values ($P_{FDR} < 0.05$), the green dots represent nominally significant *P*-values, and the grey dots represent insignificant *P*-values. The red dot indicates *PSMC3.* CN = caudate nucleus. DLPFC = dorsolateral prefrontal cortex. PCC = posterior cingulate cortex.

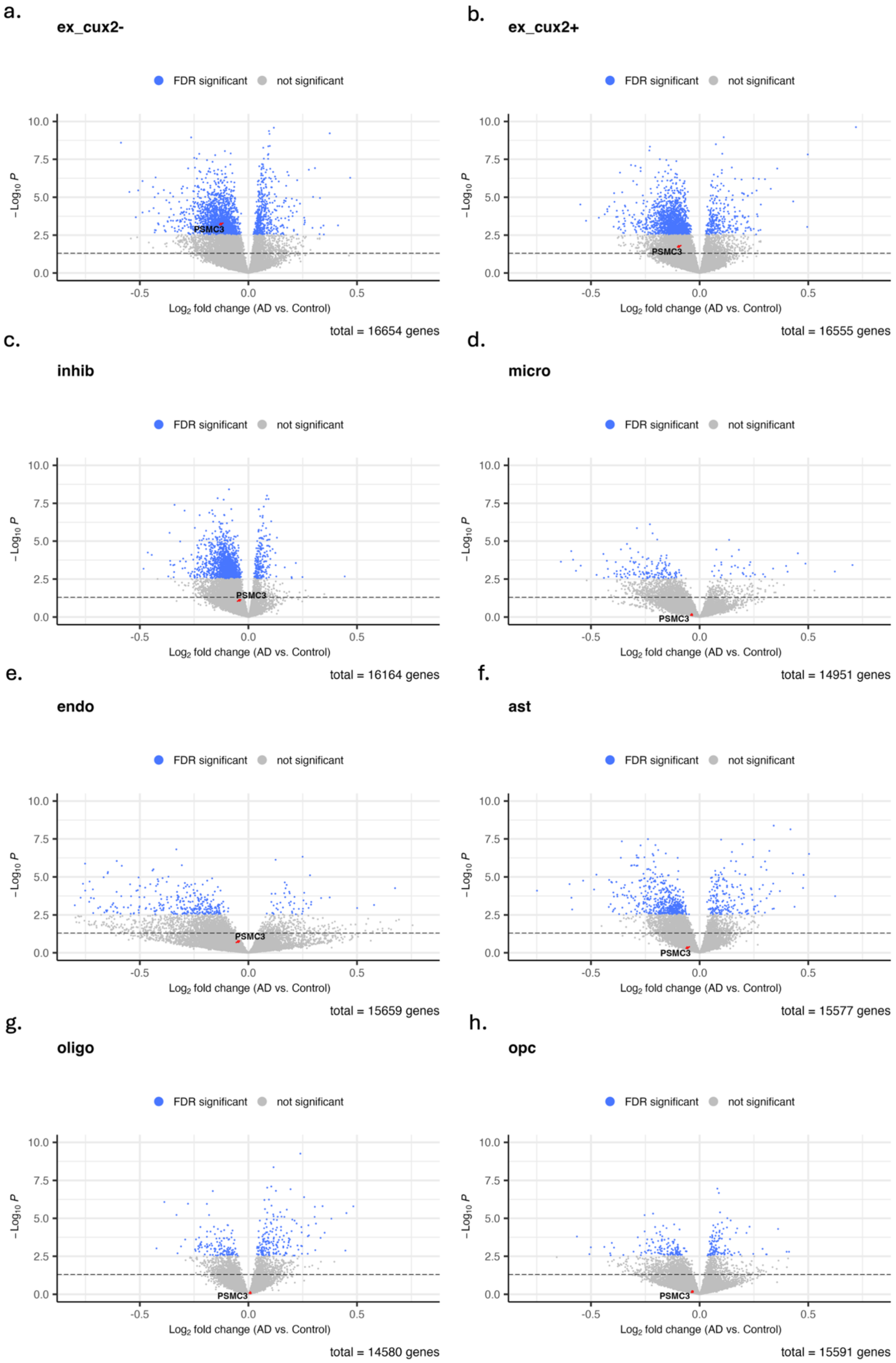

a.
ex_cux2-
FDR significant
not significant
PSMC3
total = 16654 genes
b.
ex_cux2+
FDR significant
not significant
PSMC3
total = 16555 genes
c.
inhib
FDR significant
not significant
PSMC3
total = 16164 genes
d.
micro
FDR significant
not significant
PSMC3
total = 14951 genes
e.
endo
FDR significant
not significant
PSMC3
total = 15659 genes
f.
ast
FDR significant
not significant
PSMC3
total = 15577 genes
g.
oligo
FDR significant
not significant
PSMC3
total = 14580 genes
h.
opc
FDR significant
not significant
PSMC3
total = 15591 genes
$-Log_{10}$ P
$Log_2$ fold change (AD vs. Control)

Figure S3: *PSMC3* expression levels from single-nucleus RNA sequencing in ROSMAP associated with AD case/control status across eight DLPFC cell types. The x-axis represents the log-fold change of *PSMC3* expression between AD cases and normal controls. The y-axis represents the -$\log_{10}$ *P*-values for significance. The blue dots represent FDR significant *P*-values ($P_{FDR} < 0.05$), and the grey dots represent insignificant *P*-values. The red arrow indicated *PSMC3.* ex = excitatory neurons, inhib = inhibitory neurons, micro = microglia, endo = endothelial cells, ast = astrocytes, oligo = oligodendrocytes, opc = oligodendrocyte progenitor cells.

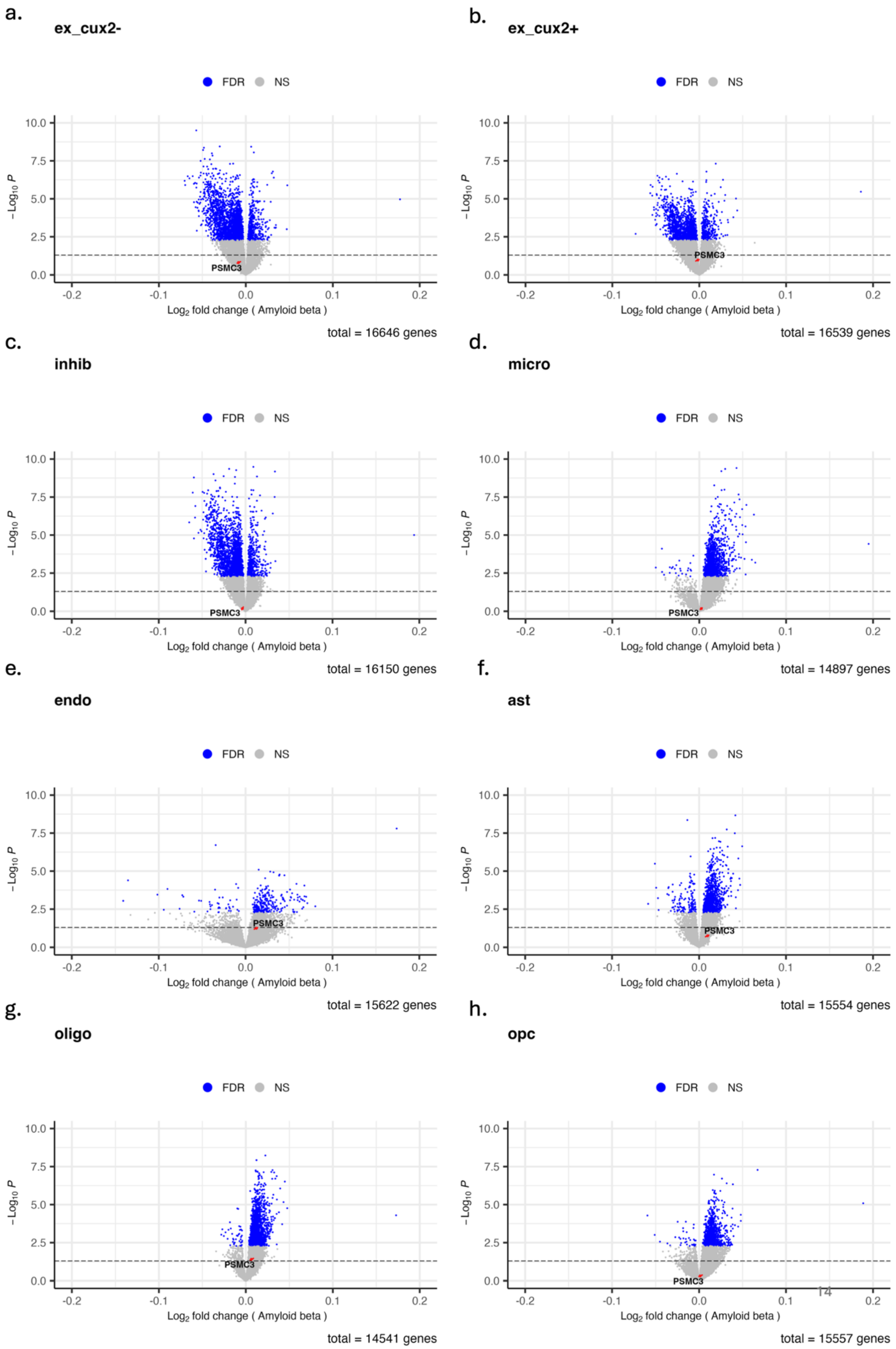
a.
ex_cux2-
FDR NS
−Log10 P
Log2 fold change ( Amyloid beta )
PSMC3
total = 16646 genes
b.
ex_cux2+
FDR NS
−Log10 P
Log2 fold change ( Amyloid beta )
PSMC3
total = 16539 genes
c.
inhib
FDR NS
−Log10 P
Log2 fold change ( Amyloid beta )
PSMC3
total = 16150 genes
d.
micro
FDR NS
−Log10 P
Log2 fold change ( Amyloid beta )
PSMC3
total = 14897 genes
e.
endo
FDR NS
−Log10 P
Log2 fold change ( Amyloid beta )
PSMC3
total = 15622 genes
f.
ast
FDR NS
−Log10 P
Log2 fold change ( Amyloid beta )
PSMC3
total = 15554 genes
g.
oligo
FDR NS
−Log10 P
Log2 fold change ( Amyloid beta )
PSMC3
total = 14541 genes
h.
opc
FDR NS
−Log10 P
Log2 fold change ( Amyloid beta )
PSMC3
total = 15557 genes

Figure S4: *PSMC3* expression levels from single-nucleus RNA sequencing in ROSMAP associated with amyloid beta across eight DLPFC cell types. The x-axis represents the log-fold change of *PSMC3* expression in relation to amyloid beta deposition. The y-axis represents the $-\log_{10}$ *P*-values for significance. The blue dots represent FDR significant *P*-values ($P_{FDR} < 0.05$), and the grey dots represent insignificant *P*-values. The red arrow indicated *PSMC3.* ex = excitatory neurons, inhib = inhibitory neurons, micro = microglia, endo = endothelial cells, ast = astrocytes, oligo = oligodendrocytes, opc = oligodendrocyte progenitor cells.

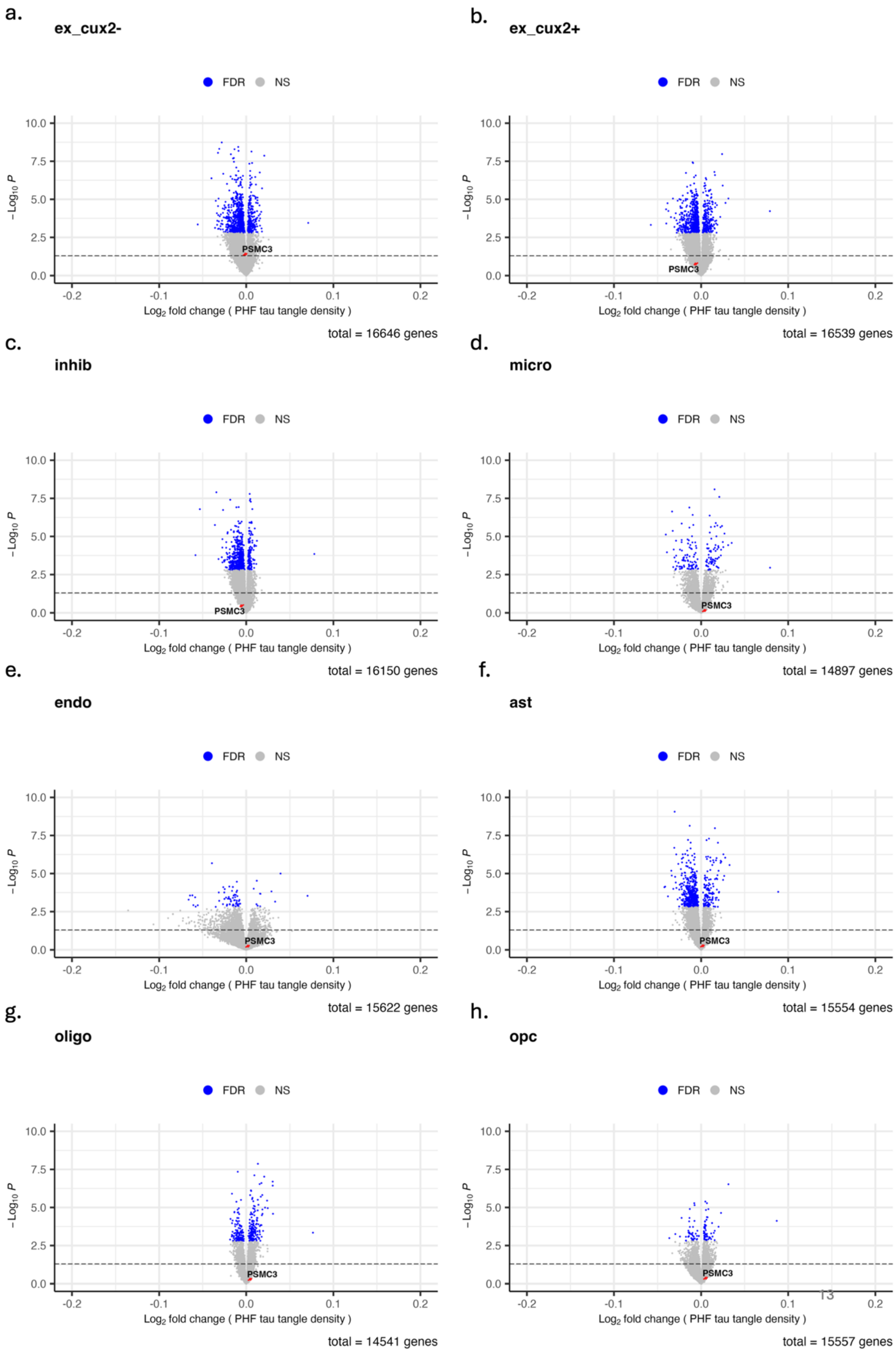
a.
ex_cux2-
FDR
NS
PSMC3
−Log10 P
Log2 fold change ( PHF tau tangle density )
total = 16646 genes
b.
ex_cux2+
FDR
NS
PSMC3
−Log10 P
Log2 fold change ( PHF tau tangle density )
total = 16539 genes
c.
inhib
FDR
NS
PSMC3
−Log10 P
Log2 fold change ( PHF tau tangle density )
total = 16150 genes
d.
micro
FDR
NS
PSMC3
−Log10 P
Log2 fold change ( PHF tau tangle density )
total = 14897 genes
e.
endo
FDR
NS
PSMC3
−Log10 P
Log2 fold change ( PHF tau tangle density )
total = 15622 genes
f.
ast
FDR
NS
PSMC3
−Log10 P
Log2 fold change ( PHF tau tangle density )
total = 15554 genes
g.
oligo
FDR
NS
PSMC3
−Log10 P
Log2 fold change ( PHF tau tangle density )
total = 14541 genes
h.
opc
FDR
NS
PSMC3
−Log10 P
Log2 fold change ( PHF tau tangle density )
total = 15557 genes

Figure S5: *PSMC3* expression levels from single-nucleus RNA sequencing in ROSMAP associated with tau tangles across eight DLPFC cell types. The x-axis represents the log-fold change of *PSMC3* expression in relation to tau tangles deposition. The y-axis represents the $-\log_{10}$ *P*-values for significance. The blue dots represent FDR significant *P*-values ($P_{FDR}$ < 0.05), and the grey dots represent insignificant *P*-values. The red arrow indicated *PSMC3.* ex = excitatory neurons, inhib = inhibitory neurons, micro = microglia, endo = endothelial cells, ast = astrocytes, oligo = oligodendrocytes, opc = oligodendrocyte progenitor cells.

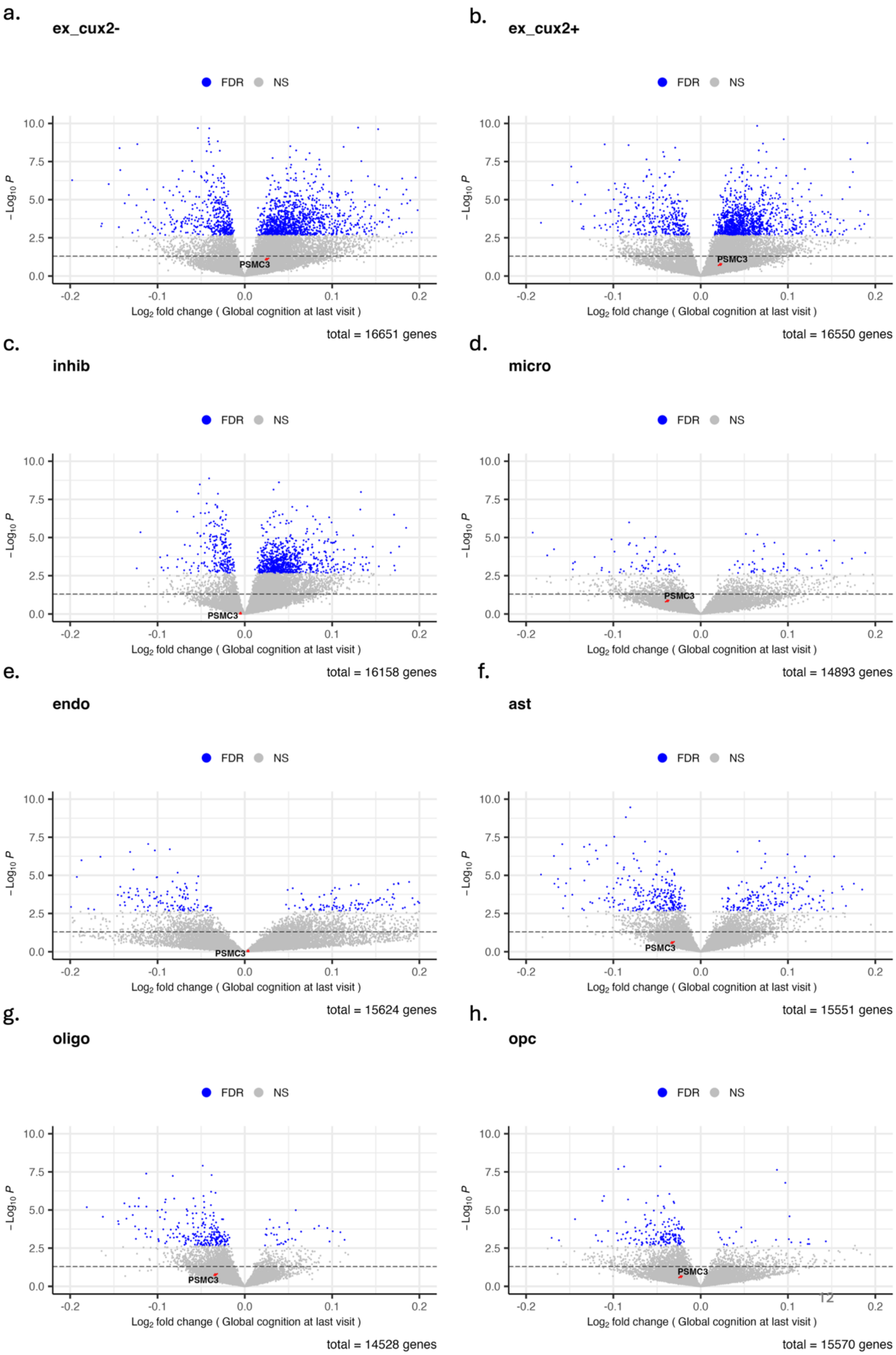
a.
ex_cux2-
FDR NS
−Log10 P
PSMC3
Log2 fold change ( Global cognition at last visit )
total = 16651 genes
b.
ex_cux2+
FDR NS
PSMC3
Log2 fold change ( Global cognition at last visit )
total = 16550 genes
c.
inhib
FDR NS
PSMC3
Log2 fold change ( Global cognition at last visit )
total = 16158 genes
d.
micro
FDR NS
PSMC3
Log2 fold change ( Global cognition at last visit )
total = 14893 genes
e.
endo
FDR NS
PSMC3
Log2 fold change ( Global cognition at last visit )
total = 15624 genes
f.
ast
FDR NS
PSMC3
Log2 fold change ( Global cognition at last visit )
total = 15551 genes
g.
oligo
FDR NS
PSMC3
Log2 fold change ( Global cognition at last visit )
total = 14528 genes
h.
opc
FDR NS
PSMC3
Log2 fold change ( Global cognition at last visit )
total = 15570 genes

Figure S6: *PSMC3* expression levels from single-nucleus RNA sequencing in ROSMAP associated with global cognition score at last visit across eight DLPFC cell types. The x-axis represents the log-fold change of *PSMC3* expression in relation to the global cognition score at the last visit. The y-axis represents the $-\log_{10}$ *P*-values for significance. The blue dots represent FDR significant *P*-values ($P_{FDR} < 0.05$), and the grey dots represent insignificant *P*-values. The red arrow indicated *PSMC3.* ex = excitatory neurons, inhib = inhibitory neurons, micro = microglia, endo = endothelial cells, ast = astrocytes*,* oligo = oligodendrocytes, opc = oligodendrocyte progenitor cells.

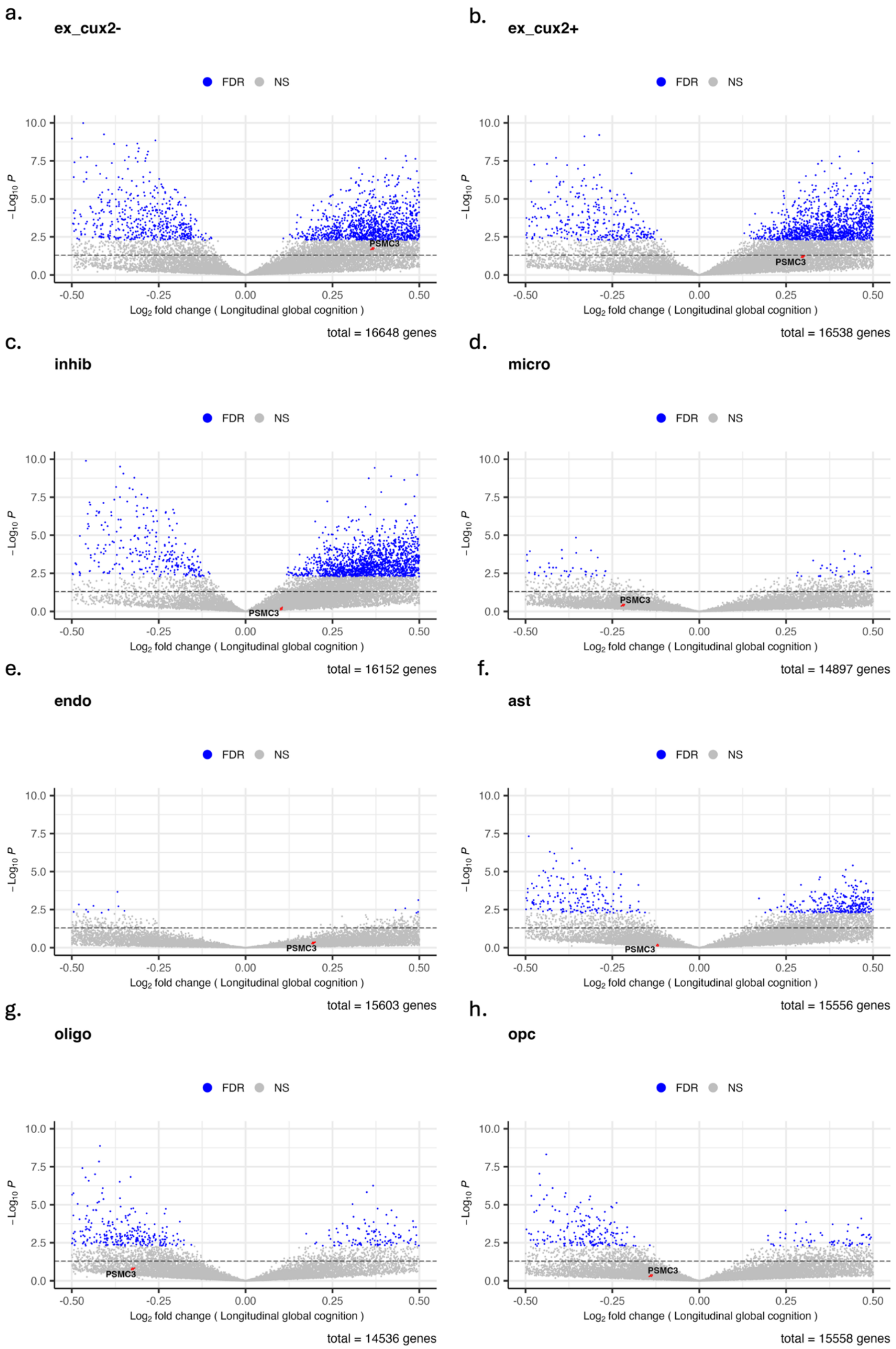

a.
ex_cux2-
FDR
NS
PSMC3
$-\log_{10} P$
$\log_2$ fold change ( Longitudinal global cognition )
total = 16648 genes
b.
ex_cux2+
FDR
NS
PSMC3
$-\log_{10} P$
$\log_2$ fold change ( Longitudinal global cognition )
total = 16538 genes
c.
inhib
FDR
NS
PSMC3
$-\log_{10} P$
$\log_2$ fold change ( Longitudinal global cognition )
total = 16152 genes
d.
micro
FDR
NS
PSMC3
$-\log_{10} P$
$\log_2$ fold change ( Longitudinal global cognition )
total = 14897 genes
e.
endo
FDR
NS
PSMC3
$-\log_{10} P$
$\log_2$ fold change ( Longitudinal global cognition )
total = 15603 genes
f.
ast
FDR
NS
PSMC3
$-\log_{10} P$
$\log_2$ fold change ( Longitudinal global cognition )
total = 15556 genes
g.
oligo
FDR
NS
PSMC3
$-\log_{10} P$
$\log_2$ fold change ( Longitudinal global cognition )
total = 14536 genes
h.
opc
FDR
NS
PSMC3
$-\log_{10} P$
$\log_2$ fold change ( Longitudinal global cognition )
total = 15558 genes

Figure S7: *PSMC3* expression levels from single-nucleus RNA sequencing in ROSMAP associated with longitudinal global cognition trajectory across eight DLPFC cell types. The x-axis represents the log-fold change of *PSMC3* expression in relation to the longitudinal global cognition trajectory. The y-axis represents the $-\log_{10}$ *P*-values for significance. The blue dots represent FDR significant *P*-values ($P_{FDR} < 0.05$), and the grey dots represent insignificant *P*-values. The red arrow indicated *PSMC3.* ex = excitatory neurons, inhib = inhibitory neurons, micro = microglia, endo = endothelial cells, ast = astrocytes, oligo = oligodendrocytes, opc = oligodendrocyte progenitor cells.

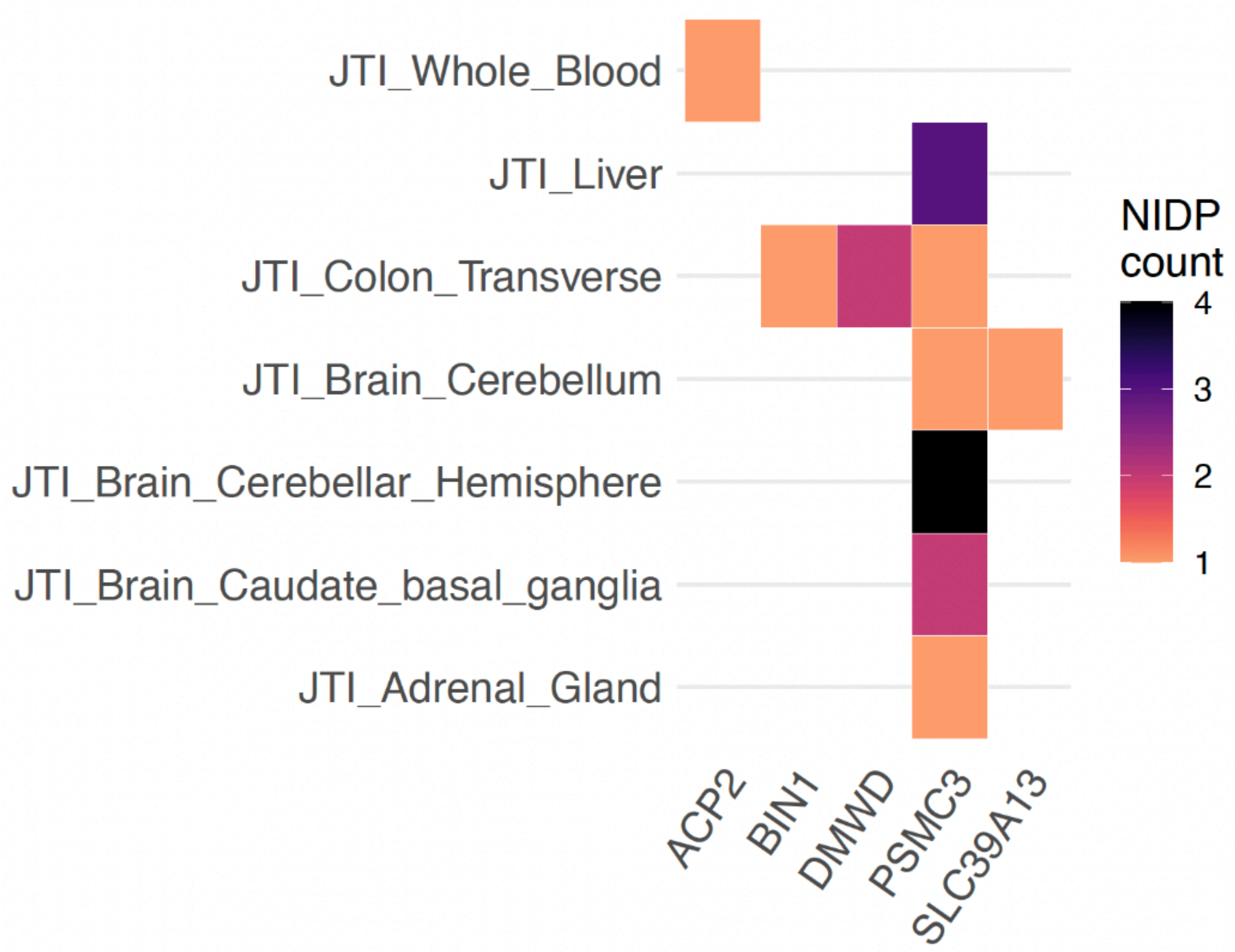


Figure S8: Tile plot detailing statistically significant associations between AD GReX measures (x-axis) and subcortical and cortical NIDPs derived from the FIRST and FAST neuroimaging MRI segmentation protocols. Associations are distributed across the tissue models in which the association was found to be significant (y-axis). These data are for the image-agnostic neuroimaging TWAS analysis. The color of each tile represents the number of NIDPs with which the gene is associated in each particular tissue model. Statistical significance was assessed via TWAS with FDR thresholding and heterogeneity adjustment applied via the MR-JTI causal inference framework. NIDP – neuroimaging derived phenotype, JTI – joint tissue imputation; MRI – magnetic resonance imaging

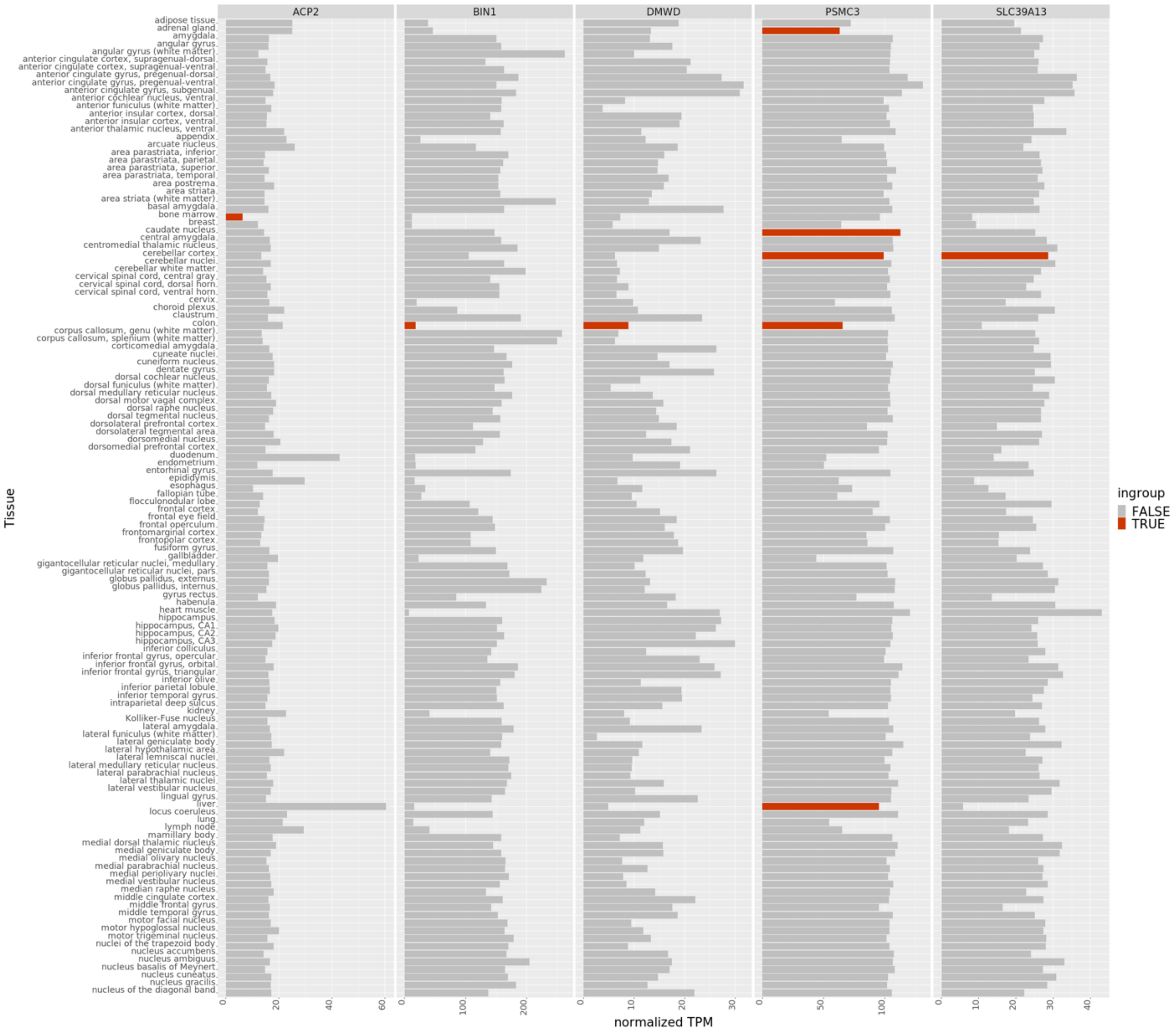


Figure S9: Expression values in normalized transcripts per million of genes associated with neuroimaging features through expression. Data reflect gene expression across all tissues in the Human Protein Atlas. Orange bars represent tissues in which the gene is significantly associated with an AD-derived neuroimaging measure. TPM – transcripts per million

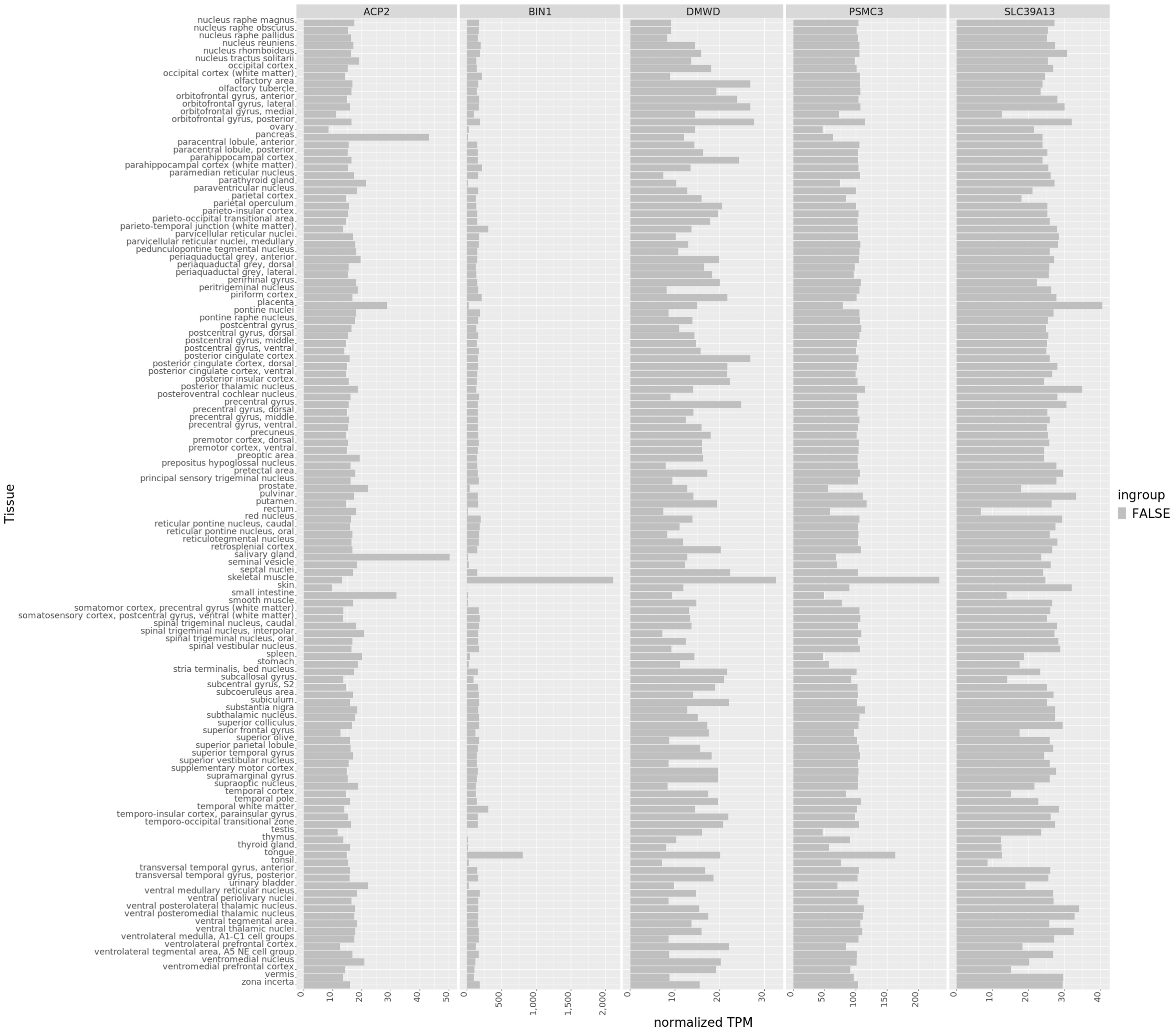


Figure S10: Expression values in normalized transcripts per million of genes associated with neuroimaging features derived from clinical AD studies through expression. Data reflect gene expression across all tissues in the Human Protein Atlas. Orange bars represent tissues in which the gene is significantly associated with an AD-derived neuroimaging measure. TPM – transcripts per million

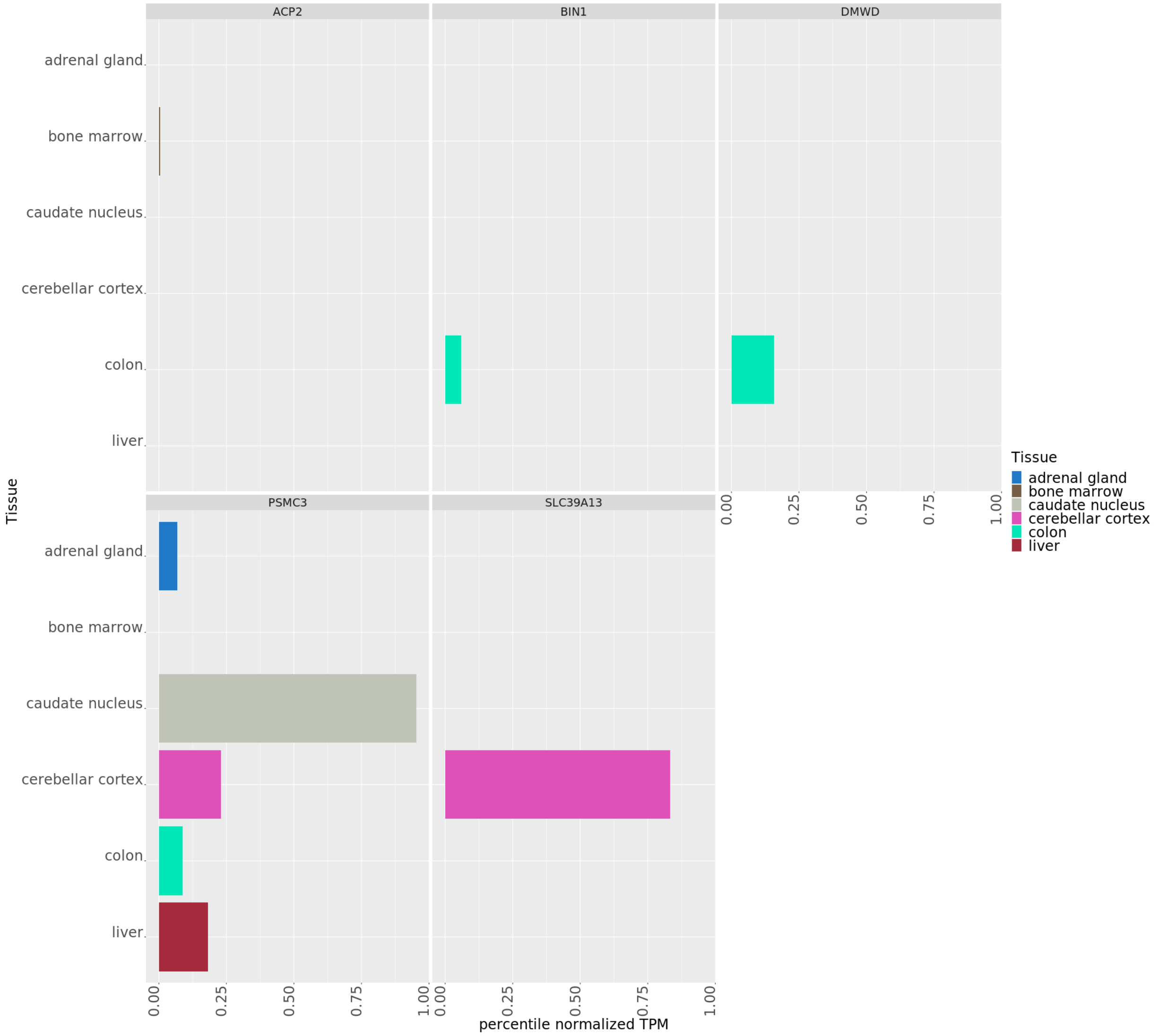


Figure S11: Tissue specific expression data of transcripts derived from the image-agnostic analysis of Alzheimer's neuroimaging data. Expression data were derived from RNA-sequencing measurements with values expressed by normalized transcripts per million (TPM) and ranked by percentile.

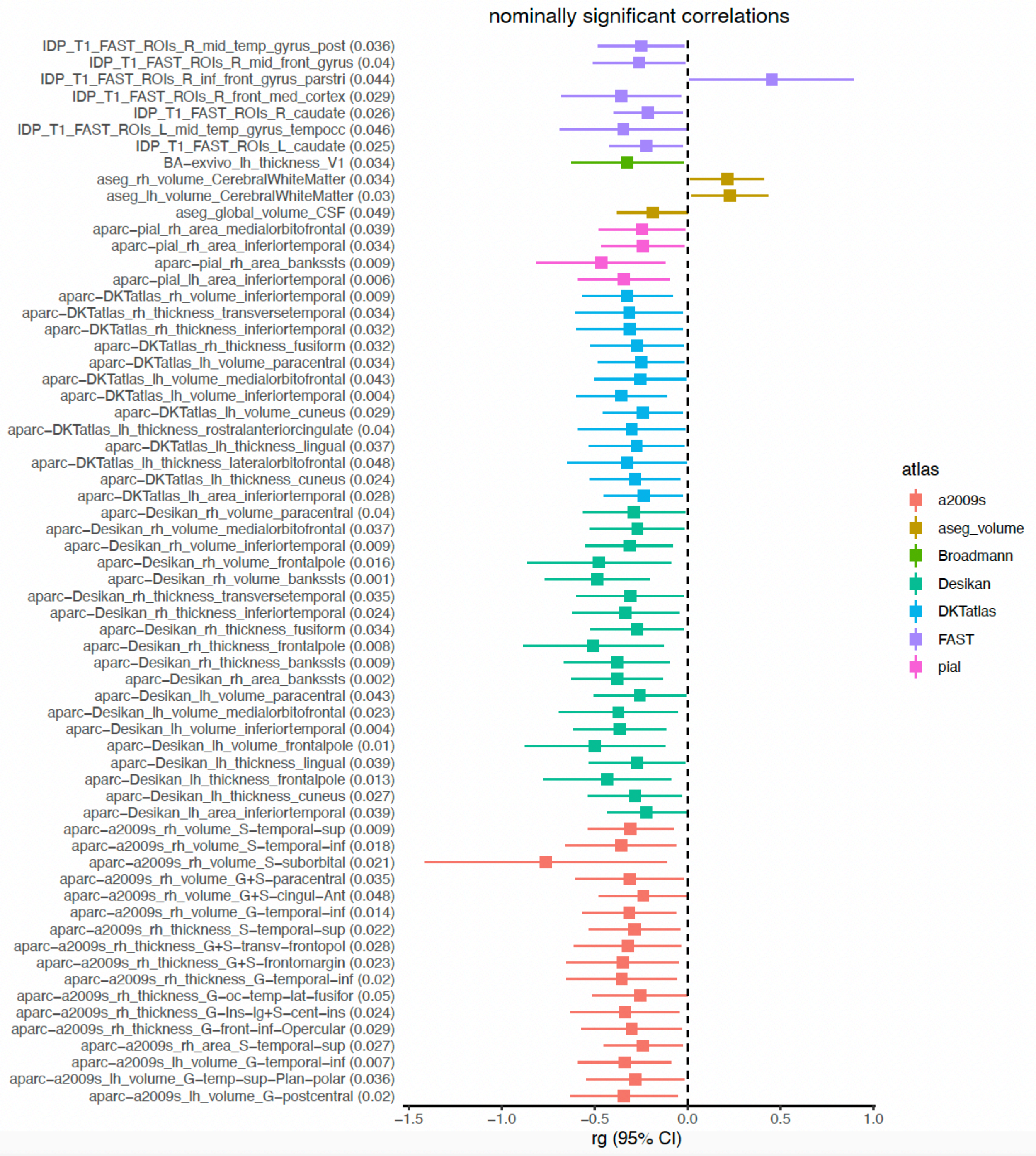


Figure S12: Nominally significant global genetic correlations between NIDP and family history of dementia across all T1-derived NIDPs. Genetic correlation was calculated using linkage disequilibrium score regression correlation analysis.

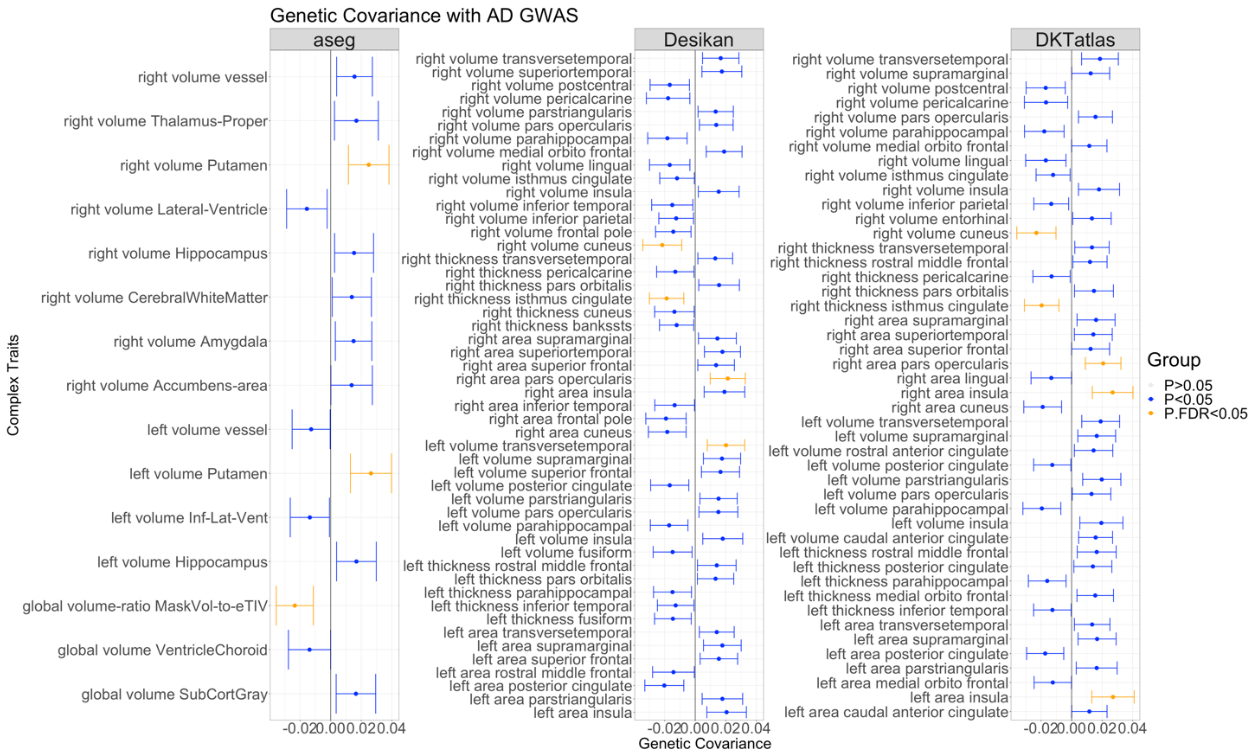


Figure S13: Genetic covariance results performed using GeNetic cOVariance Analyzer (GNOVA) between Neuroimaging Derived Phenotypes (NIDPs), processed using aseg, Desikan, and DKT atlases, and clinical Alzheimer's Disease status indicated by Kunkle et al.'s study. The x-axis represents the genetic covariance estimates. The y-axis demonstrates the NIDPs. Each facet represents the results from different atlas. The FDR-significant results are colored orange, and the nominally significant results are colored blue. The non-significant results are not shown here. FDR – false discovery rate; NIDP – neuroimaging-derived phenotypes

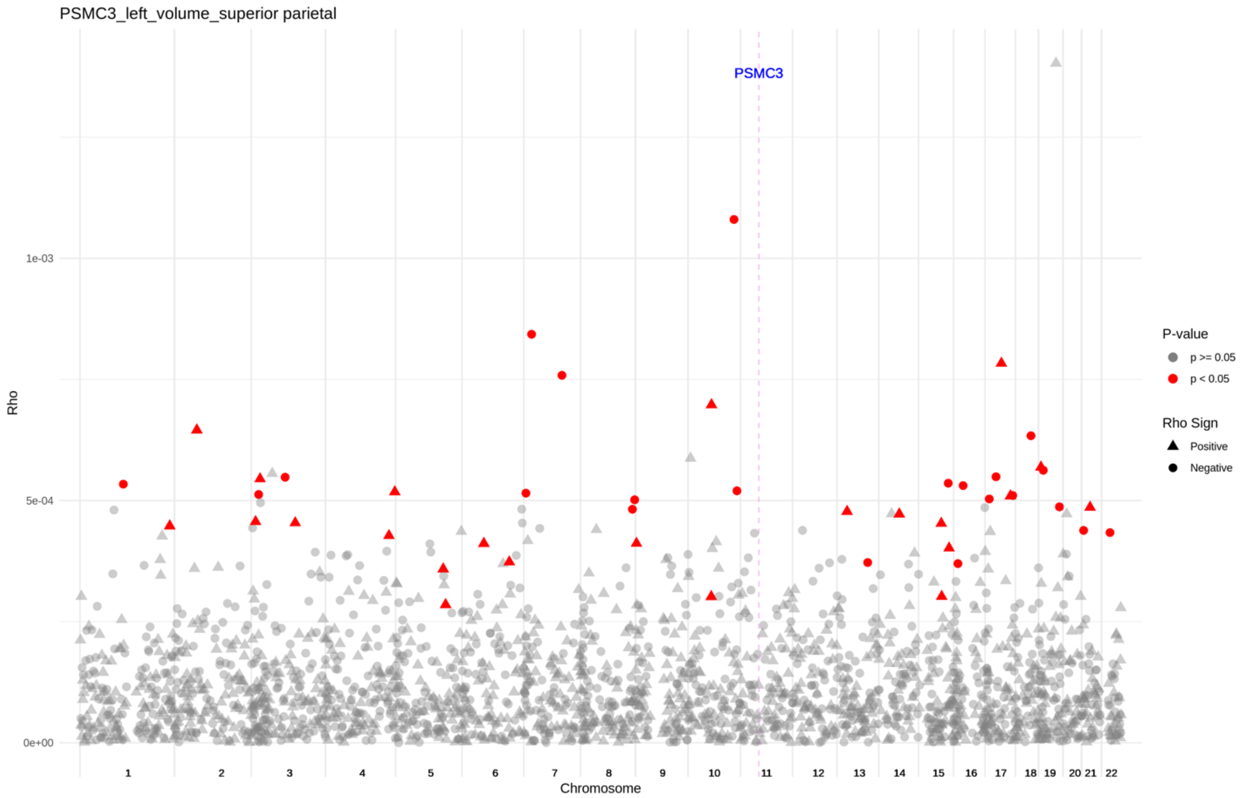


Figure S14: Genome-wide results from the local genetic covariance analysis between the family history of dementia GWAS by Marioni et al. and the listed NIDP from the image-directed analysis result prior to the Mendelian randomization (MR) test. Each point represents a single locus, with the y-axis location indicating the covariance between SNPs effect sizes in that region. The color of the point represents significance with red indicating nominal significance. As each NIDP was identified according to associations with AD-genes, we overlayed the location of the associated gene onto the superGNOVA findings. Significant loci occurring within 2 megabase of the gene of interest are emphasized with larger size.

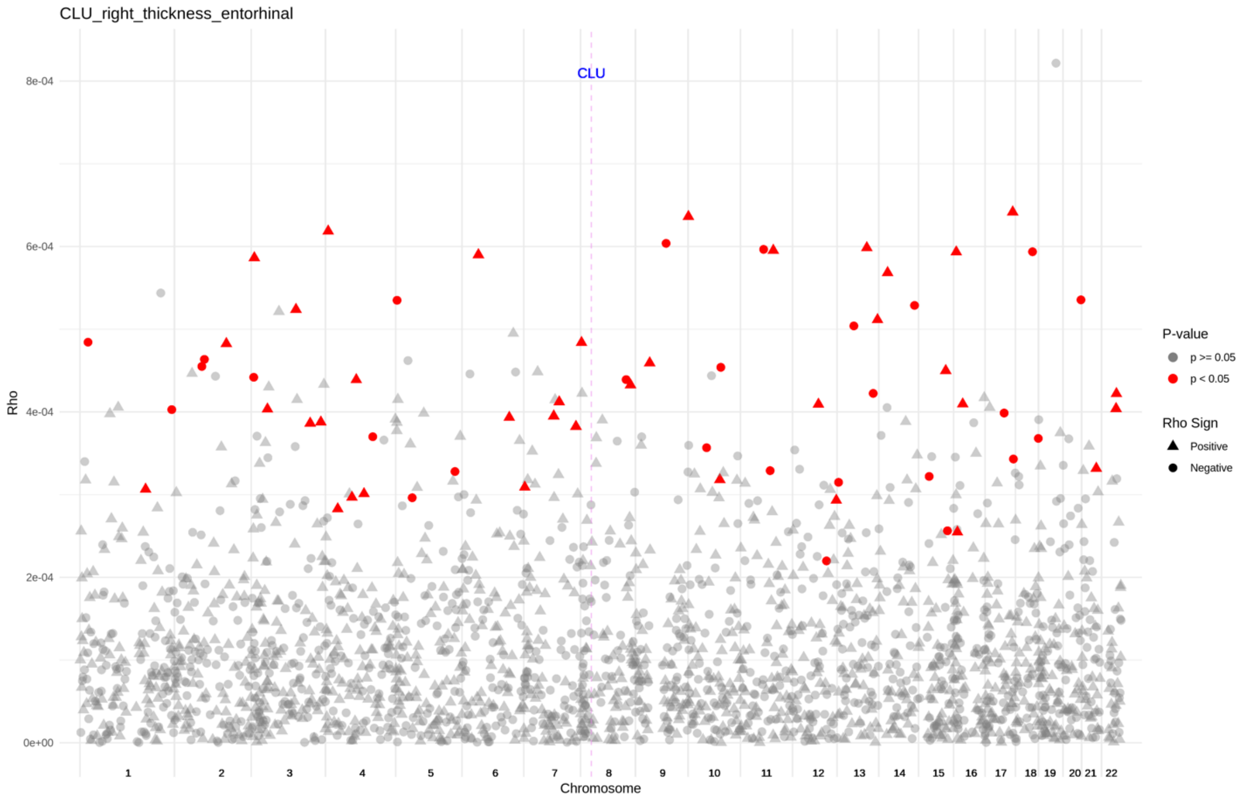


Figure S15: Genome-wide results from the local genetic covariance analysis between the family history of dementia GWAS by Marioni et al. and the listed NIDP from the image-directed analysis result prior to the MR test. Each point represents a single locus with the y-axis location indicating the covariance between SNPs effect sizes in that region. The color of the point represents significance with red indicating nominal significance. As each NIDP was identified according to associations with AD-genes, we overlayed the location of the associated gene onto the superGNOVA findings. Significant loci occurring within 2 megabase of the gene of interest are emphasized with larger size.

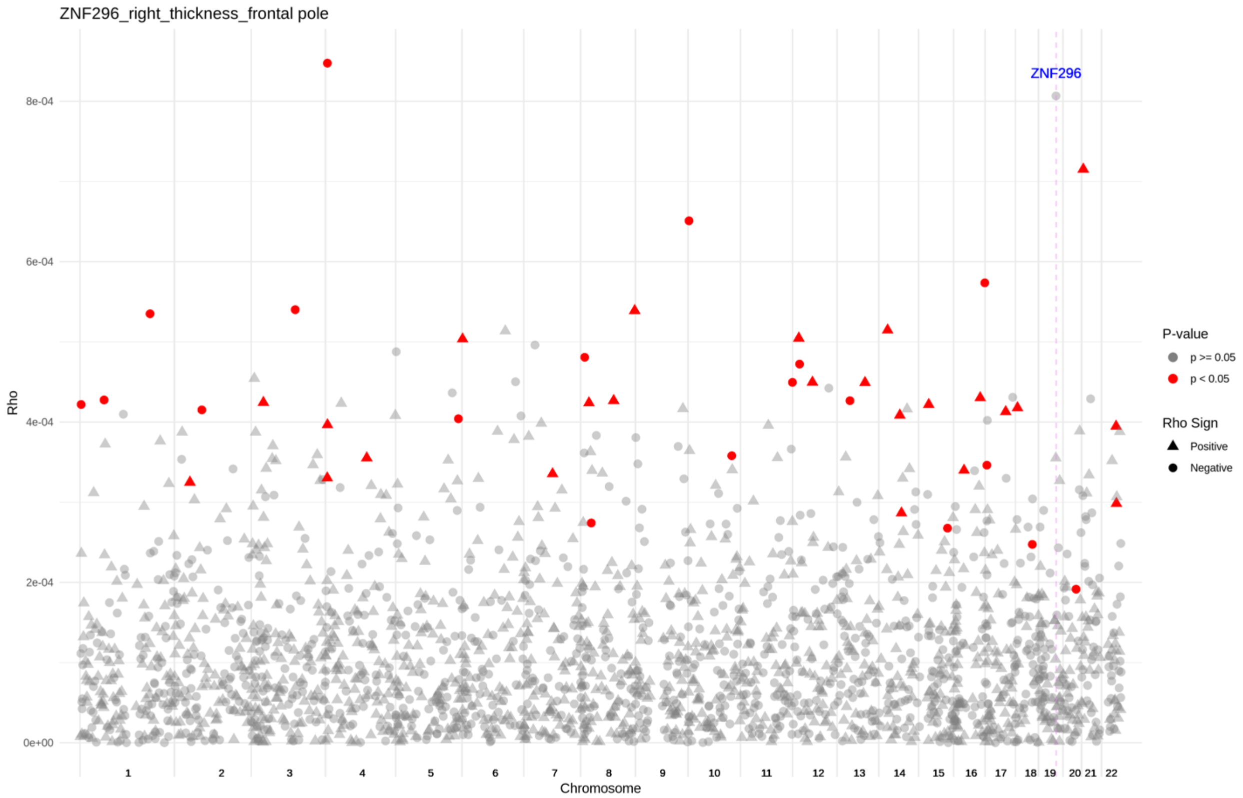


Figure S16: Genome-wide results from the local genetic covariance analysis between the family history of dementia GWAS by Marioni et al. and the listed NIDP from the image-directed analysis result prior to the MR test. Each point represents a single locus with the y-axis location indicating the covariance between SNPs effect sizes in that region. The color of the point represents significance with red indicating nominal significance. As each NIDP was identified according to associations with AD-genes, we overlayed the location of the associated gene onto the superGNOVA findings. Significant loci occurring within 2 megabase of the gene of interest are emphasized with larger size.

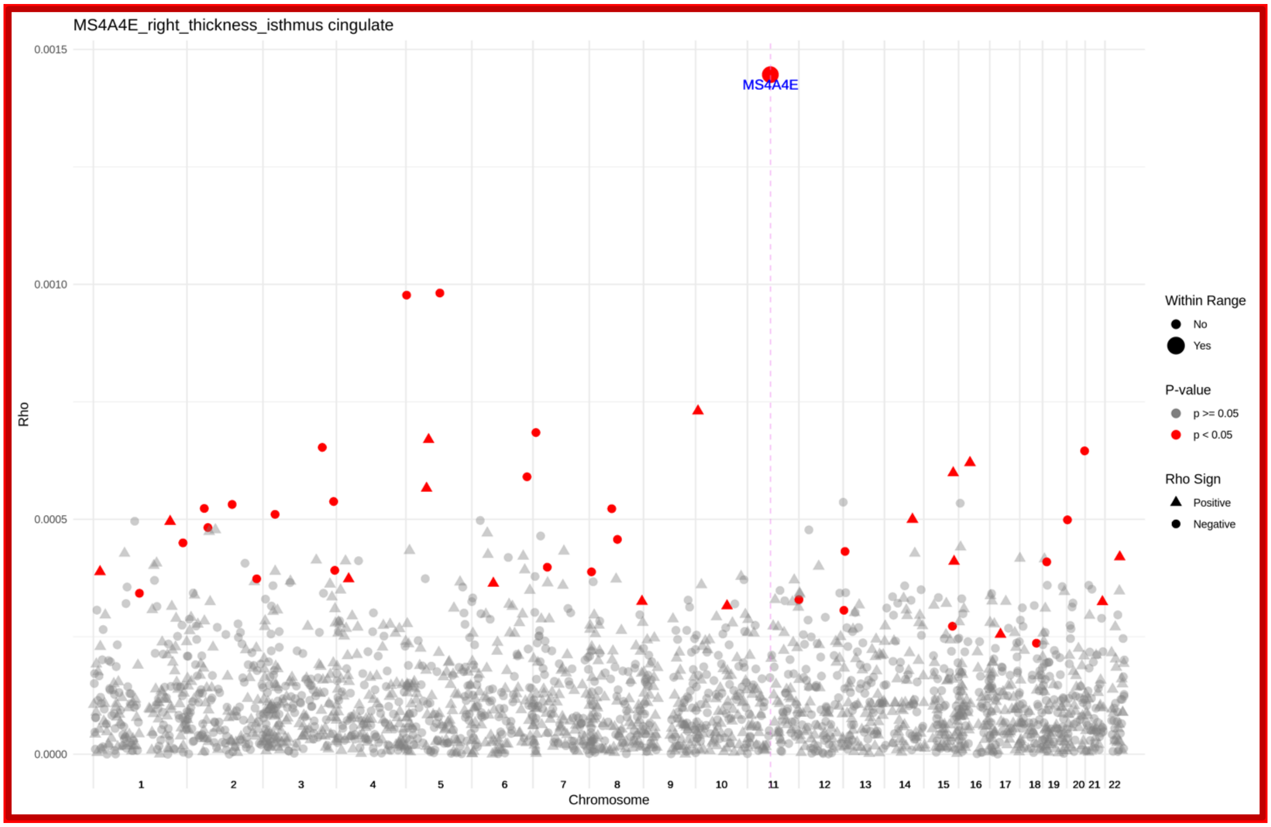


Figure S17: Genome-wide results from the local genetic covariance analysis between the family history of dementia GWAS by Marioni et al. and the listed NIDP from the image-directed analysis result prior to the MR test. Each point represents a single locus with the y-axis location indicating the covariance between SNPs effect sizes in that region. The color of the point represents significance with red indicating nominal significance. As each NIDP was identified according to associations with AD-genes, we overlayed the location of the associated gene onto the superGNOVA findings. Significant loci occurring within 2 megabase of the gene of interest are emphasized with larger size.

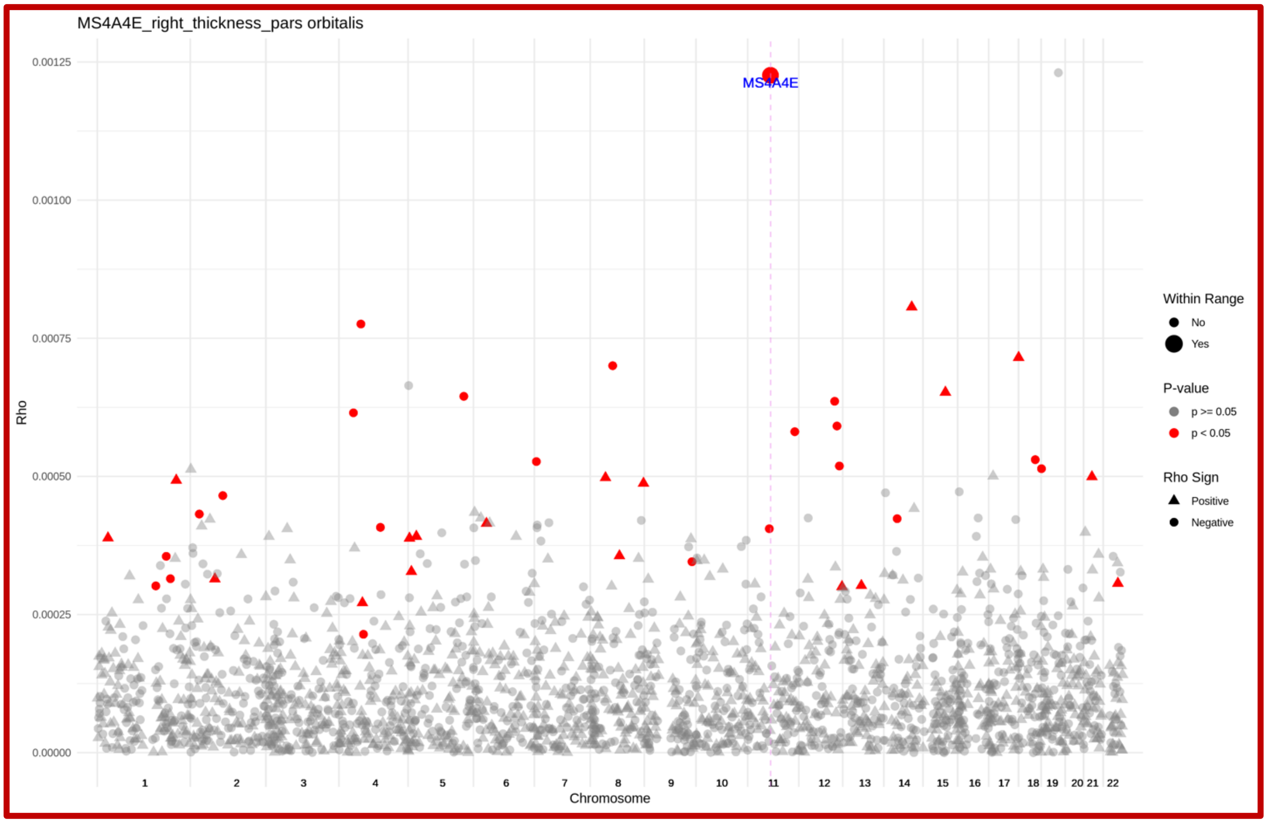


Figure S18: Genome-wide results from the local genetic covariance analysis between the family history of dementia GWAS by Marioni et al. and the listed NIDP from the image-directed analysis result prior to the MR test. Each point represents a single locus with the y-axis location indicating the covariance between SNPs effect sizes in that region. The color of the point represents significance with red indicating nominal significance. As each NIDP was identified according to associations with AD-genes, we overlayed the location of the associated gene onto the superGNOVA findings. Significant loci occurring within 2 megabase of the gene of interest are emphasized with larger size.

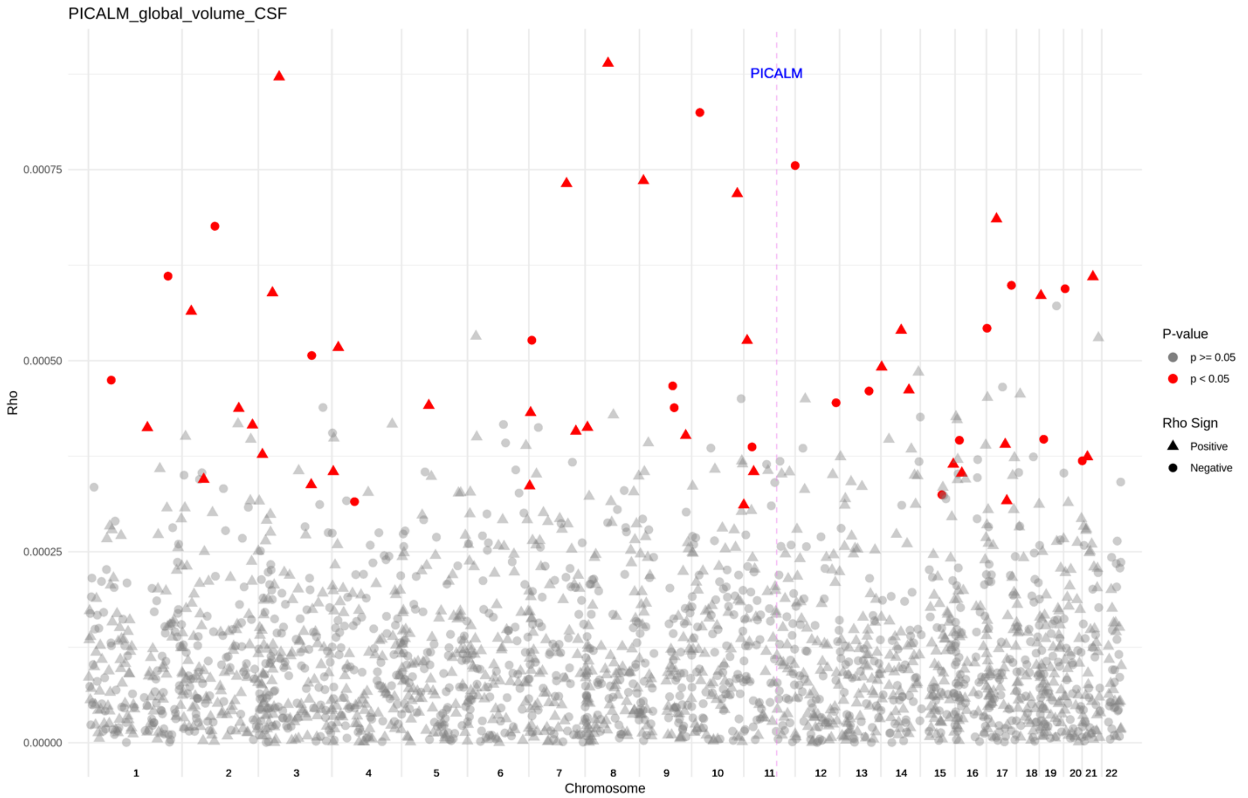


Figure S19: Genome-wide results from the local genetic covariance analysis between the family history of dementia GWAS by Marioni et al. and the listed NIDP from the image-directed analysis result prior to the MR test. Each point represents a single locus with the y-axis location indicating the covariance between SNPs effect sizes in that region. The color of the point represents significance with red indicating nominal significance. As each NIDP was identified according to associations with AD-genes, we overlayed the location of the associated gene onto the superGNOVA findings. Significant loci occurring within 2 megabase of the gene of interest are emphasized with larger size.

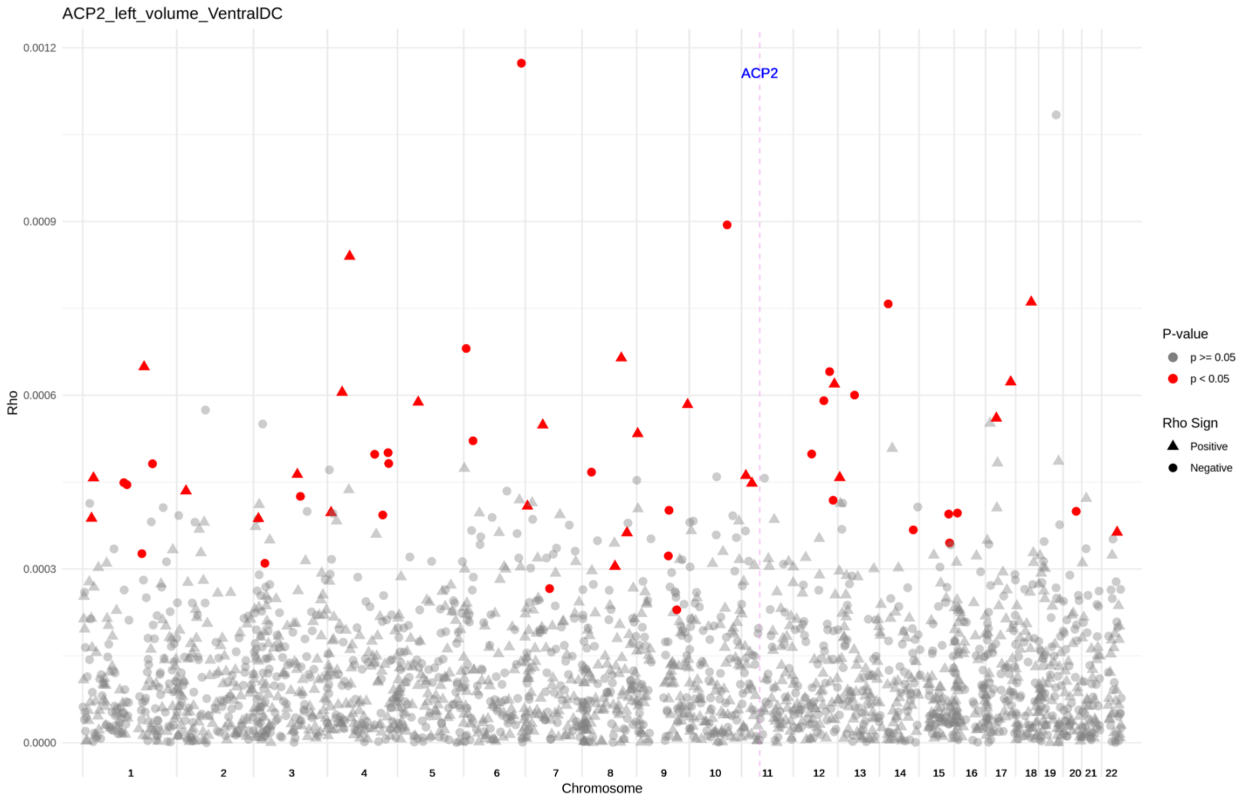


Figure S20: Genome-wide results from the local genetic covariance analysis between the family history of dementia GWAS by Marioni et al. and the listed NIDP from the image-directed analysis result prior to the MR test. Each point represents a single locus with the y-axis location indicating the covariance between SNPs effect sizes in that region. The color of the point represents significance with red indicating nominal significance. As each NIDP was identified according to associations with AD-genes, we overlayed the location of the associated gene onto the superGNOVA findings. Significant loci occurring within 2 megabase of the gene of interest are emphasized with larger size.

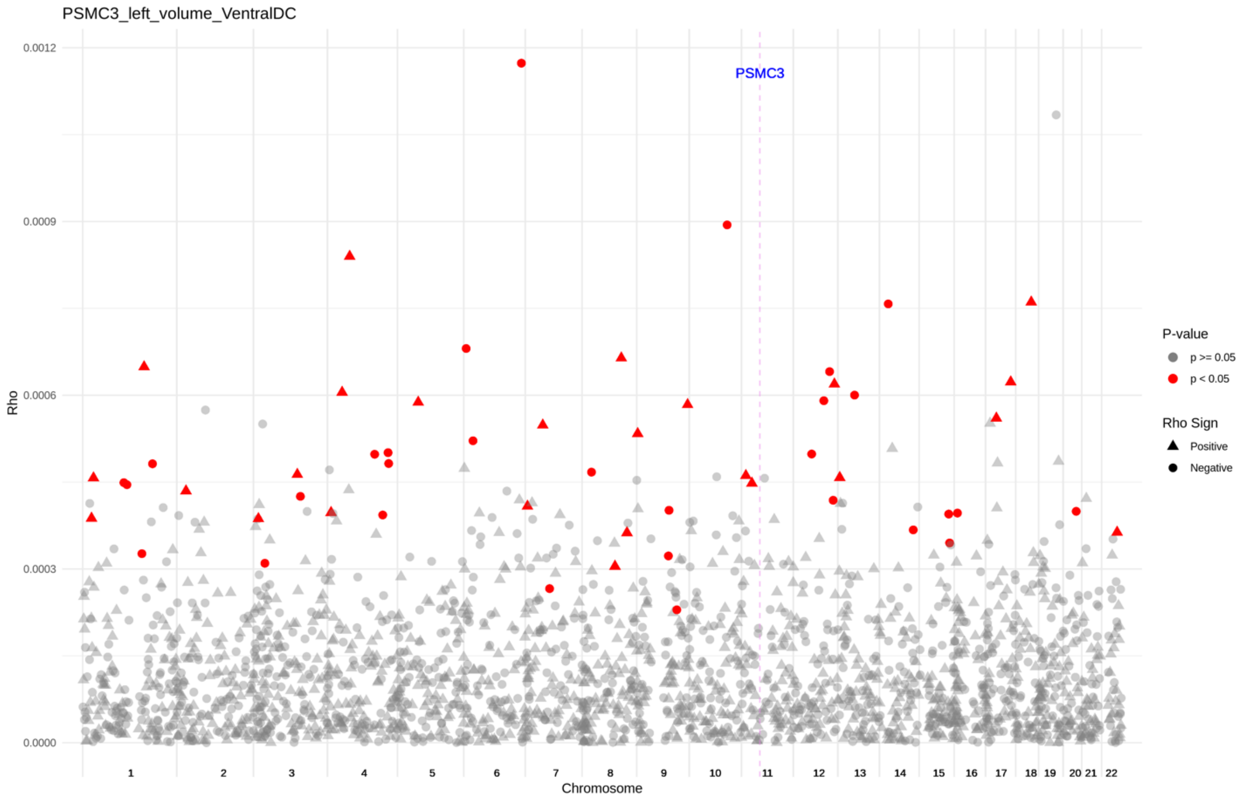


Figure S21: Genome-wide results from the local genetic covariance analysis between the family history of dementia GWAS by Marioni et al. and the listed NIDP from the image-directed analysis result prior to the MR test. Each point represents a single locus with the y-axis location indicating the covariance between SNPs effect sizes in that region. The color of the point represents significance with red indicating nominal significance. As each NIDP was identified according to associations with AD-genes, we overlayed the location of the associated gene onto the superGNOVA findings. Significant loci occurring within 2 megabase of the gene of interest are emphasized with larger size.

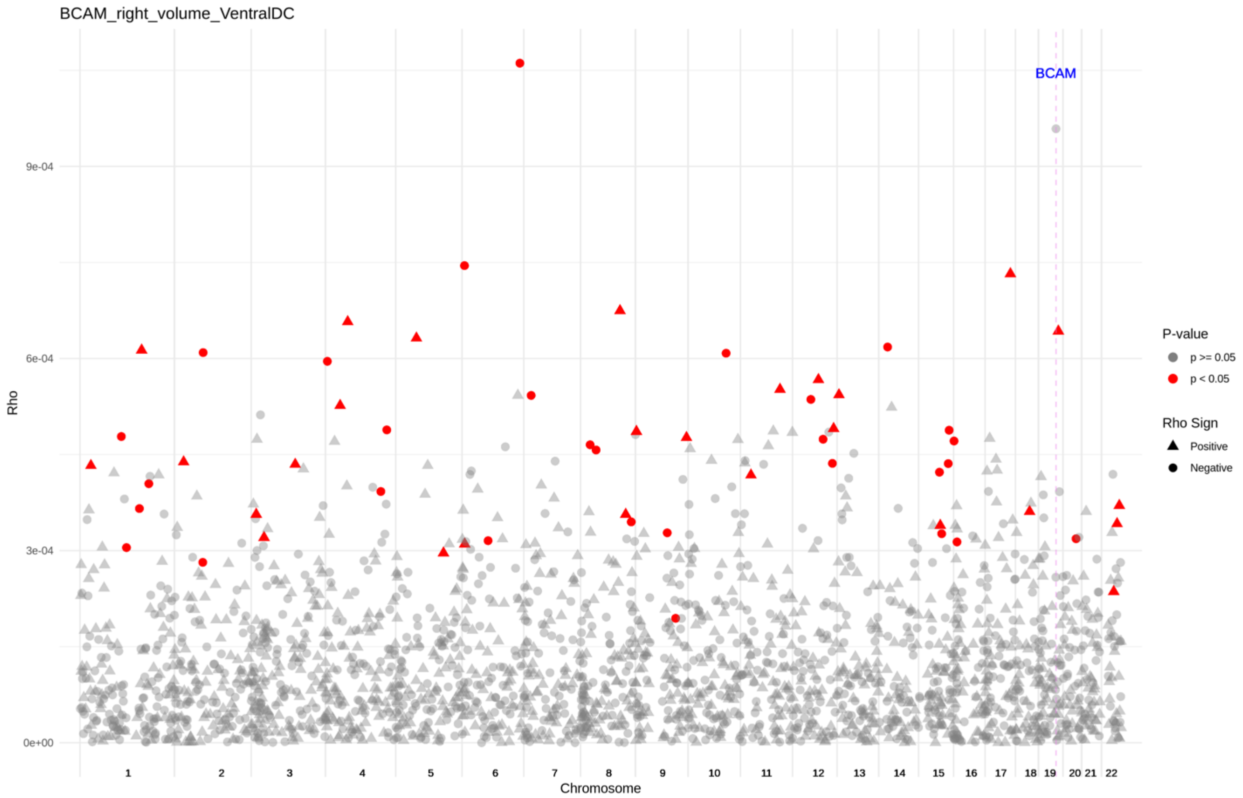


Figure S22: Genome-wide results from the local genetic covariance analysis between the family history of dementia GWAS by Marioni et al. and the listed NIDP from the image-directed analysis result prior to the MR test. Each point represents a single locus with the y-axis location indicating the covariance between SNPs effect sizes in that region. The color of the point represents significance with red indicating nominal significance. As each NIDP was identified according to associations with AD-genes, we overlayed the location of the associated gene onto the superGNOVA findings. Significant loci occurring within 2 megabase of the gene of interest are emphasized with larger size.

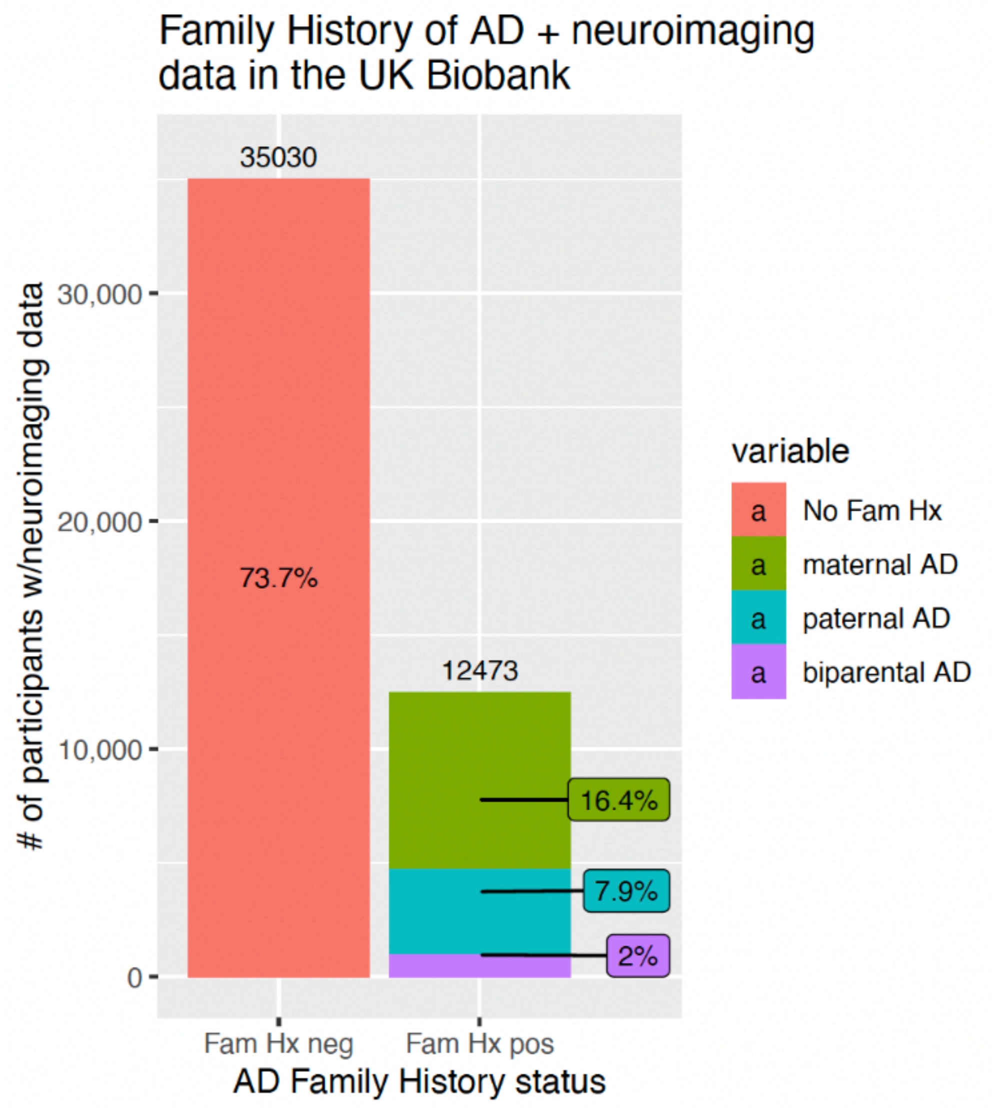


Figure S23: Distribution of individuals in the UK Biobank with and without self-reported parental history of Alzheimer's disease and related dementias, with positive cases split according to the parent. Fam Hx – family history; AD – Alzheimer's disease
